# I3LUNG: Clinical Validation of a Multimodal AI Tool to Support Immunotherapy Decisions in NSCLC

**DOI:** 10.64898/2026.01.16.25342913

**Authors:** Arsela Prelaj, Vanja Miskovic, Matteo Sacco, Alberto Ferrarin, Cristina Licciardello, Leonardo Provenzano, Margherita Favali, Ludovica Lerma, Aleksandra Zec, Andrea Spagnoletti, Monica Ganzinelli, Daniele Lorenzini, Beshoy Guirges, Luca Invernizzi, Cecilia Silvestri, Laura Mazzeo, Marco Meazza Prina, Giulia Corrao, Margherita Ruggirello, Andra Diana Dumitrascu, Rosa Di Mauro, Dario Monzani, Gabriella Pravettoni, Michele Zanitti, Davide Macocchi, Moreno Bruno Marino, Chiara Cavalli, Rebecca Romanò, Claudia Giani, Samuel G. Armato, Alessandra Esposito, Christine Bestvina, Maria Spector, Bogot R Naama, Reham Basheer, Adi Lahiani Hafzadi, Laila Roisman, Iris Watermann, Marlen Szewczyk, Till Olchers, Heinz Richter, Constantin Blanke-Roeser, Costanza Siniscalchi, Nikolaos Spathas, Evangelos Sarris, Elena Fountzilas, Aina Arbusà Roca, Rocio Caro-Consuegra, Patricia Iranzo, Melissa Fernández-Pinto, Jose Rodríguez-Morató, Luca Agnelli, Mario Occhipinti, Marta Brambilla, Teresa Beninato, Claudia Proto, Sokol Kosta, Michele Pio Di Palma, Eliana Rulli, Stefan Steurer, Ronald Simon, Michael Willis, Giancarlo Pruneri, Filippo De Braud, Marcello Restelli, Enriqueta Felip, Nir Peled, Alexander T Pearson, Helena Linardou, Martin Reck, Giuseppe Lo Russo, Francesco Trovò, Alessandra Pedrocchi, Marina Chiara Garassino

## Abstract

Despite a decade of immunotherapy, treatment selection in non-small cell lung cancer (NSCLC) still relies on subgroup analyses and clinical scores. I3LUNG (NCT05537922) is currently the largest international, real-world, multimodal, artificial intelligence (AI)-based trial, enrolling 2365 patients. We integrated real-world clinical data (RWD), computed tomography (CT) images, digital pathology (DP), and genomics (G) into machine learning early-fusion (MLEF) and deep-learning intermediate-fusion (DLIF) models. MLEF achieved consistent performance across outcomes (AUC≈0.74), with improved results in first-line patients (AUC up to 0.82). Multimodal models outperformed RWD in clinical-specific subgroups (AUCs up to 0.86). In the test set, AI models surpassed PD-L1, ECOG PS, NLR, LDH (all with *p*<0.01) and the LIPI score. The clinical usability study showed that expert and non-expert physicians could improve their prediction with the explainable AI (XAI) tool. The I3LUNG tool emerges as a clinically relevant decision-support system and is currently under prospective validation in >2,000 patients.

## Introduction

Immunotherapy (IO) targeting programmed cell death protein 1 (PD-1)^1–3^, programmed death-ligand 1 (PD-L1)^4^, and cytotoxic T-lymphocyte associated protein 4 (CTLA-4)^5^, have transformed metastatic non-small cell lung cancer (NSCLC) care, achieving long-term benefit in 20-30% of patients^6^. IO alone or in combination with chemotherapy (IO/CHT) currently forms the backbone of advanced non oncogene-addicted NSCLC treatment; yet, most patients face primary (5–20%) or secondary (60–85%) resistance.^7^ This underscores the need for robust predictive biomarkers to identify likely IO responders at diagnosis, avoiding unnecessary toxicity and cost. Despite its limited predictive power, PD-L1 remains the only clinically-approved biomarker.^8^

Beyond traditional methods, artificial intelligence (AI)-based approaches leverage large-scale, complex data to enable individualized IO outcome prediction. While unimodal studies (e.g., genomics^9^, radiomics^10^, digital pathology^11^) show promise, they remain limited by size, scope, and single-center design.^12^ Two recent studies demonstrated the potential of multimodal integration for predicting immunotherapy response in NSCLC. Vanguri *et al.*^13^ combined CT, histopathology, and genomics, while Captier *et al.*^14^ integrated PET, pathology, transcriptomics, and clinical data. While both models outperformed traditional biomarkers, they were limited by small cohorts (≈250-300 patients), with only ≈ 80 patients having complete multimodal data.

To support individualized IO decisions, the I3LUNG (NCT05537922) project^15^ was launched under Horizon Europe 2021-2027, as an international, multicentric study. It includes retrospective/prospective data to develop and validate AI-based physician decision support systems (PDSSs) for optimizing IO treatment selection in advanced NSCLC. While prospective patient enrollment is ongoing, this study focuses on the retrospective arm of the I3LUNG project (***Supplementary Figure 1.***). We describe the multimodal data collection pipeline and present the first set of AI-based PDSSs developed using data from patients with advanced NSCLC treated with immunotherapy (IO) or chemo-immunotherapy (IO/CHT) between September 2012 and October 2023. These patients were enrolled across six clinical centers, including four in the EU (INT-Italy, MH-Greece, GHD-Germany, VHIO-Spain) and two outside the EU (UOC-USA, SZMC-Israel). We introduce a multimodal framework for personalized treatment selection comprising: foundation model-based extraction from medical images, comparative evaluation of early and intermediate fusion models, explainability and fairness analyses, and clinical usability studiy assessing real-world utility.

## Results

### Overview of the Study Design

The overview of this study is shown in **Fig. 1a**. As of January 15, 2025, 2396 stage IIIC–IVB advanced NSCLC patients treated with IO or IO/CHT were enrolled from the six clinical centers. The dataset included clinical real-world data (RWD, n=2396), genomics (G, n=1723), CT scans (n=1705) analyzed via PyRadiomics (PYRAD) and foundation models (FMRAD), and histopathology slides (DP, n=936) analyzed via foundation models. The consort flow for all data modalities is described in ***Extended data Fig.1***. Patients were allocated into three cohorts: C1: stage III NSCLC patients who received IO as maintenance treatment after chemoradiation therapy; C2: stage IV NSCLC patients who receive IO as first-line treatment for metastatic disease; and C3: stage IV NSCLC patients who receive IO as second-or further-line therapy. The present analysis focused on C2 and C3 (C2, n=1437, C3, n=638). Clinical endpoints included real-world overall survival (OS), survival status at 6 months (OS6) and 24 months (OS24), and disease control rate (DCR).

**Figure 1.**
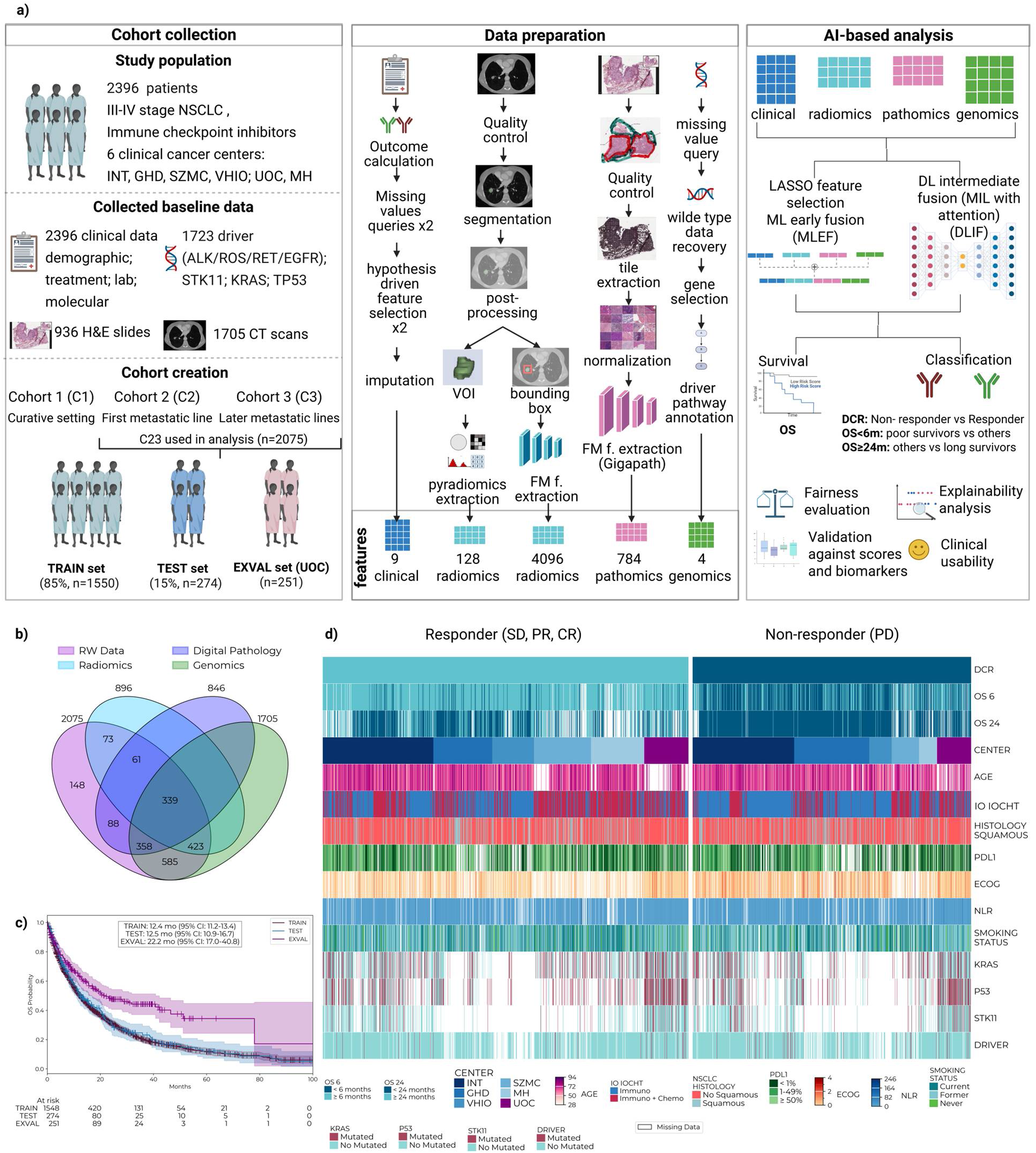
Overview of the study and dataset description. a) Study overview. Cohort collection: four data modalities from NSCLC patients included in the overall study cohort (left panel); Data preparation: showing different workflows for data pre-processing and feature extraction for each modality, including RWD, RAD (two pipelines), DP and G (middle panel); and AI-based analysis: overview of two different approaches, MLEF and DLIF, and performed analyses (right panel). b) Cohort C23 modality overview (n=2075 patients), 339 patients had all four modalities available; c) Kaplan-Meier survival curves for TRAIN, TEST and EXVAL (UOC), using OS as the endpoint; d) Multimodal cohort heatmap listing characteristics for n=2075 patients. White indicates missing information. *FM-Foundation Model, ML-Machine Learning, DL-Deep Learning, MIL-Multiple Instance Learning, OS-Overall Survival, DCR-Disease Control Rate*

The main predictive AI model was built using the combined cohort C23 (C2 and C3, n=2075). Patients from UOC served as an external validation (EXVAL, n=251) cohort to assess generalizability on cross-continental data. The remaining 1824 patients were split into a training set (TRAIN, 85%, n=1550) with 5-fold cross-validation (CV, using centers as folds) and a test set (TEST, 15%, n=274). Subgroup analyses were conducted by training and evaluating additional submodels using clinically-relevant subgroups as input: C2 only; PD-L1 ≥50% and <50%; patients who received IO as monotherapy; patients who received IO/CHT; patients with squamous cell carcinoma and adenocarcinoma histology tumors; single centers.

We used two AI-approaches for the multimodal integration: machine learning early fusion (MLEF) and deep learning intermediate fusion models (DLIF). Survival models, including Cox, were built to achieve OS outcome predictions, while classification models were used for OS6, OS24 and DCR predictions. Finally, model post-hoc explainability (XAI) analysis, fairness evaluation and a clinical usability study were performed on the TEST to assess the models’ clinical relevance and trustworthiness.

### Clinical characteristics of patients included in the analysis and multimodal data description

**Table 1** shows patient characteristics (C23). Median OS was 12.4 months (95%CI 11.2-13.4) for TRAIN, 12.5 months (95%CI 10.9-16.7) for TEST, and 22.2 months (95%CI 17.0-40.8) for EXVAL. At data cut-off, 9.3% of patients remained on treatment, and 29.8% of patients were alive (**Fig. 1c**). **Fig. 1d** presents the heatmap of the main baseline characteristics, single biomarkers, and OS6 and OS24 among DCR responders (stable disease, SD; partial response, PR; complete response, CR) vs non-responders (progressive disease, PD). For C23, RWD and both images (RAD and DP) modalities were available for 400 patients, while 339 patients had data from all the four modalities (RWD, RAD, DP and G) (**Fig. 1b**).

**Table 1.** Patients’ characteristics of datasets used in the study: TRAIN, TEST and EXVAL. *Clinical usability set is composed of subsets of TEST (n=80) and EXVAL (n=20)

|  | **TRAIN set**<br>**(n=1550)** | **TEST set**<br>**(n=274)** | **EXVAL set**<br>**(n=251)** | **Clinical usability set\***<br>**(n=100)** |
| --- | --- | --- | --- | --- |
| **Age, median (range)** | 68 (28-94) | 67 (30-87) | 69 (36-92) | 67 (30-87) |
| **Sex, n (%)** |  |  |  |  |
| Male | 1013 (65.3) | 175 (63.9) | 103 (41.0) | 60 (60) |
| Female | 537 (34.6) | 99 (36.1) | 148 (59.0) | 40 (40) |
| NA | 0 (0) | 0 (0) | 0 (0) | 0 (0) |
| **ECOG Performance Status, n (%)** |  |  |  |  |
| ECOG 0 | 470 (30.3) | 90 (32.9) | 6 (2.4) | 27 (27) |
| ECOG 1 | 725 (46.8) | 115 (42) | 197 (78.5) | 48 (48) |
| ECOG $\geq 2$ | 194 (12.5) | 43 (14.9) | 45 (18) | 16 (16) |
| NA | 161 (10.4) | 26 (9.5) | 3 (1.2) | 9 (9) |
| **Histology, n (%)** |  |  |  |  |
| Adenocarcinoma | 1005 (64.8) | 178 (65) | 207 (82.5) | 74 (74) |
| Squamous | 367 (23.7) | 70 (25.6) | 27 (10.8) | 17 (17) |
| Other | 162 (10.5) | 23 (8.4) | 13 (5.2) | 9 (9) |
| NA | 16 (1) | 3 (1) | 4 (1.5) | 0 (0) |
| **Smoking status, n (%)** |  |  |  |  |
| Current | 537 (34.7) | 101 (36.9) | 39 (15.5) | 27 (27) |
| Former | 754 (48.7) | 133 (48.5) | 189 (75.3) | 58 (58) |
| Never | 117 (7.6) | 19 (7) | 23 (9.2) | 6 (6) |
| NA | 142 (9.2) | 21 (7.7) | 0 (0) | 9 (9) |
| **PD-L1 expression, n (%)** |  |  |  |  |
| <1% | 428 (27.6) | 75 (27.4) | 101 (40.2) | 30 (30) |
| 1-49% | 383 (24.7) | 74 (27) | 50 (19.9) | 28 (28) |
| $\geq 50\%$ | 424 (27.4) | 66 (24.1) | 69 (27.5) | 30 (30) |
| NA | 315 (20.3) | 59 (21.5) | 31 (12.4) | 12 (12) |
| **Therapy type, n (%)** |  |  |  |  |
| IO | 904 (58.3) | 161 (58.8) | 96 (38.2) | 50 (50) |
| IO/CHT | 646 (41.7) | 113 (41.2) | 155 (61.8) | 50 (50) |
| NA | 0 (0) | 0 (0) | 0 (0) | 0 (0) |
| **Therapy line, n (%)** |  |  |  |  |
| 1 (c2) | 1046 (67.5) | 181 (66.1) | 210 (83.7) | 96 (96) |
| 2 (c3) | 344 (22.2) | 66 (24.1) | 36 (14.3) | 4 (4) |
| ≥ 3 (c3) | 160 (10.3) | 27 (9.8) | 5 (2.0) | 0 (0) |
| NA | 0 (0) | 0 (0) | 0 (0) | 0 (0) |
| **Disease control rate, n (%)** |  |  |  |  |
| PD | 681 (43.9) | 104 (38.1) | 109 (43.4) | 35 (35) |
| SD/PR/CR | 863 (55.7) | 169 (61.9) | 142 (56.6) | 65 (65) |
| NA | 6 (0.4) | 1 (0.4) | 0 (0) | 0 (0) |
| **Survival months, median) (95%CI)** |  |  |  |  |
| **OS** | 12.4 (11.2-13.4) | 12.5 (10.9-16.7) | 22.2 (17.0-40.8) | 13.6 (11.1-20.9) |
| **PFS** | 5.0 (4.5-5.4) | 5.1 (4.3-5.8) | 8.0 (6.0-11.1) | 6.1 (4.2-9.8) |

To develop predictive tools, we trained and evaluated MLEF and DLIF using multiple modality combinations, and different clinically-uniform patients subgroups, such as first-line IO patients and those with PD-L1 ≥ 50% (full list of analysis in *Supplementary Table 1*).

### Machine learning-based multimodal integration predicts IO effectiveness

Since the MLEF workflow requires complete data, patients with missing modalities were excluded, resulting in varying sample sizes for each analysis **(Fig. 2a)**. To ensure a fair comparison, we compared them with the corresponding RWD-only models on the same subset of patients (RWD-only matched). Multimodal MLEF models were compared on the CV, due to the small TEST size. In general, the main analysis RWD-only models provided strong baseline performance across endpoints in CV (**Fig. 2ci-iii**), achieving AUC of 0.74 (n=226; 95% CI: 0.67–0.81) in TEST, and 0.60 (n=190; 95% CI: 0.52–0.68) in EXVAL for OS24; AUC 0.69 (n=259; 95% CI: 0.62–0.76) in TEST, and 0.64 (n=240; 95% CI: 0.56–0.72) in EXVAL for OS6; and of 0.71 (n=273; 95% CI: 0.65–0.77) in TEST, and 0.55 (n=251; 95% CI: 0.48–0.62) in EXVAL for DCR. Among unimodal analyses, FMRAD features stood out for predicting OS24, (CV AUC 0.72, 95% CI: 0.67–0.77), while other modalities alone, excluding RWD, contributed modestly, with AUC lower than 0.65, **Fig. 2b**.

**Figure 2.**
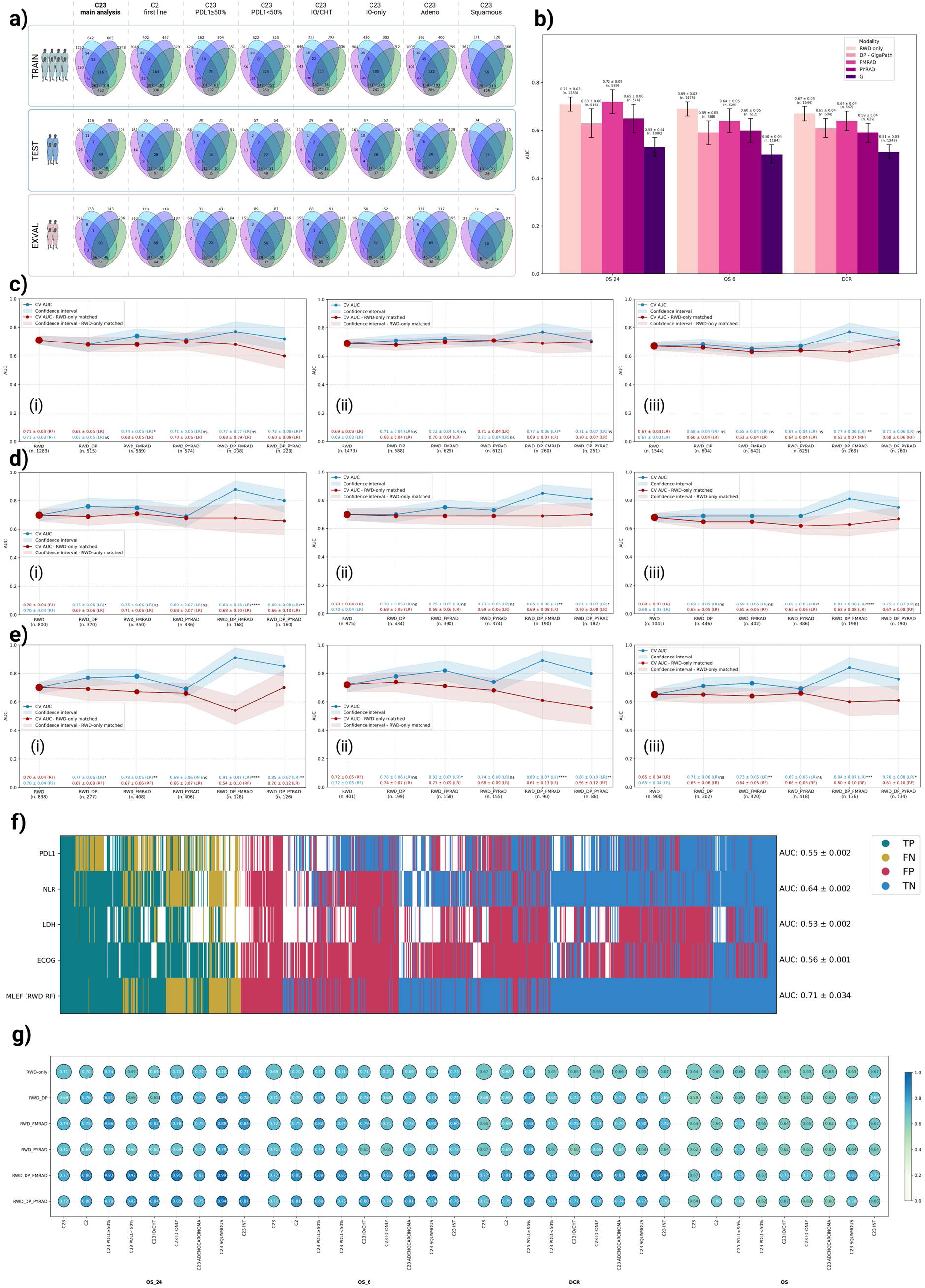
Machine Learning Early Fusion (MLEF) framework: a) MLEF subanalysis with number of pts in TRAIN; TEST and EXVAL for each subanalysis; b) Unimodal MLEF performance across outcomes; c) Classification performance of MLEF unimodal, bimodal, and multimodal models for three endpoints: (i) OS24; (ii) OS6; (iii) DCR, evaluated on the C23 cohort, cross-validation (CV); Multimodal model performance (blue) was compared with RWD-only model on the same dataset (RWD-only matched - red); performance assessed using AUC and tested with DeLong’s method, p values calculated using two-sided t-test, n.s. - not significant, * p≤0.05, ** p≤0.01, *** p≤0.001, **** p≤0.0001; Classification performance of MLEF unimodal, bimodal, and multimodal models for three endpoints: (i) OS24; (ii) OS6; (iii) DCR, evaluated on the C2 cohort, cross-validation (CV); performance assessed using AUC and tested with DeLong’s method, p values calculated using two-sided t-test, n.s. - not significant, * p≤0.05, ** p≤0.01, *** p≤00..001, **** p≤0.0001; d) Selected MLEF sub-analyses for classification: (i) OS24 in the IO-only cohort, (ii) OS6 in the PD-L1 ≥50% cohort, (iii) DCR in the IO-only cohort; performance assessed using AUC and tested with DeLong’s method, p values calculated using two-sided t-test, n.s. - not significant, * p≤0.05, ** p≤0.01, *** p≤0.001, **** p≤0.0001; e) Patient-level comparison of MLEF and single biomarkers (C23) in CV for predicting long responders (OS24): each row represents one patient, ordered from left (mostly correctly predicted positives) to right (mostly correctly predicted negatives), with the middle section highlighting patients where models and biomarkers consistently struggled; f) CV AUC performance of MLEF classifier; (i) OS24, (ii) OS6, (iii) DCR and Cox-MLEF, (iv) C-index across all subgroup analysis; circle color indicates the model’s performance, while size corresponds to the size of pts cohort. *C23-cohort 23, C2-Cohort 2, TP-True Positive, TN-True Negative, FP-False Positive, FN-False Negative, OS-Overall Survival, DCR-Disease Control Rate, NLR - Neutrophil-to-Lymphocyte Ratio*

Specific combinations of modalities led to the improved performance across all three classification endpoints (OS24, OS6, and DCR, **Fig. 2ci-iii**), with the best results observed when RWD was integrated with DP and RAD (**Fig. 2c and 2g**). The added value of multimodality was particularly evident in predicting DCR, where RWD+DP+FMRAD models showed significant improvement vs RWD-only matched models (CV AUC 0.77 (95% CI: 0.71–0.83) vs 0.63 (95% CI: 0.56–0.70), *p*≤0.01), **Fig. 2ciii**. Similarly, for OS24 prediction, RWD+DP+PYRAD performed better than RWD-only matched (CV AUC: 0.72 (95% CI: 0.64–0.80) vs 0.60 (95% CI: 0.51–0.69), *p*≤0.05), **Fig. 2ci**. Patient case-level comparisons for OS24 (**Fig. 2f**) demonstrated that the MLEF RWD-only random forest (RF) model provided a rate of correct predictions more balanced across the two classes as compared to single biomarkers.

Similarly, in the first-line setting (C2) the results improved with multimodal integration, **Fig. 2di-iii**. RWD+DP+FMRAD outperformed RWD-only matched: for instance, the addition of DP and FMRAD improved AUC from 0.68 (95% CI: 0.58-0.78) to 0.88 (95% CI: 0.82-0.94, *p*≤0.0001) for OS24, and from 0.63 (95% CI: 0.55-0.71) to 0.81 (95% CI: 0.75-0.87, *p*≤0.0001) for DCR.

### Survival analysis achieved moderate results

For the combined C23 analysis, the full RWD-only model achieved a C-index of 0.64 (95% CI: 0.62–0.66) in CV, 0.66 (95% CI: 0.62–0.70) in TEST, and 0.65 (95% CI: 0.60–0.70) in EXVAL. Similarly, in the C2 analysis, the RWD-only model achieved a C-index of 0.65 (95% CI: 0.63–0.67) in CV, 0.66 (95% CI: 0.60–0.72) in TEST, and 0.66 (95% CI: 0.61–0.71) in EXVAL. In both cohorts, the multimodal model with RWD+DP+FMRAD outperformed the RWD-only matched model in CV (C23: 0.67 (95% CI: 0.63–0.71) vs 0.61 (95% CI: 0.57–0.65), *p*≤0.05; C2: 0.74 (95% CI: 0.70–0.78) vs 0.63 (95% CI: 0.58–0.68), *p*≤0.00001). Full C-index results are reported in ***Extended Data Fig 2d***.

### Multimodal approaches outperformed RWD-only in NSCLC-specific subpopulations

For OS24, multimodal approaches outperformed RWD-only matched models in IO-only patients (AUC 0.91 vs 0.54, *p*≤0.0001, **Fig. 2ei**) and other subgroups including PD-L1≥50%, IO/CHT, adenocarcinoma, squamous, and INT-only **(*Extended Data Fig.3a*)**. Similar trends were observed for OS6 (e.g., PD-L1≥50%, AUC 0.89 vs 0.61, *p*≤0.0001, **Fig. 2eii**) and DCR (e.g., IO-only, AUC 0.84 vs 0.60, *p*≤0.001, **Fig. 2diii**). For OS Cox models, improvements were seen in PD-L1≥50%, squamous, and IO-only groups. However, multimodal models did not outperform RWD-only matched models in all subanalysis. A summary of CV AUCs across all endpoints and subgroups is shown in **Fig. 2g**, with full metrics reported in ***Extended Data Fig.3***.

### Deep learning framework did not outperform machine learning one

The DLIF approach (**Fig.3a**), which retains all patients regardless of missing modalities, achieved similar performance to MLEF with RWD-only (AUC for OS24: CV 0.68 (95% CI: 0.65–0.71), TEST 0.73 (95% CI: 0.66–0.80), EXVAL 0.63 (95% CI: 0.55–0.71); OS6: CV 0.69, (95% CI: 0.66–0.72), TEST 0.72 (95% CI: 0.65–0.79), EXVAL 0.72, 95% CI: 0.64–0.80); DCR: CV 0.69 (95% CI: 0.67–0.71), TEST 0.70 (95% CI: 0.64–0.76), EXVAL 0.65 (95% CI: 0.58–0.72), **Fig. 3bi-iii**. Regardless of the outcome, the DLIF approach did not benefit from adding other modalities. This held across endpoints and in subgroup analyses, including first-line treatment (C2), **Fig. 3c and *Extended Data Fig. 3***. Similarly, Cox-DLIF showed no advantage over MLEF (C-index: CV 0.64 (95% CI: 0.62–0.66), TEST: 0.65 (95% CI: 0.60–0.70), EXVAL 0.63 (95% CI: 0.58–0.68)), with no benefit from multimodal integration *(**Extended Data Fig. 3**)*.

**Figure 3.**
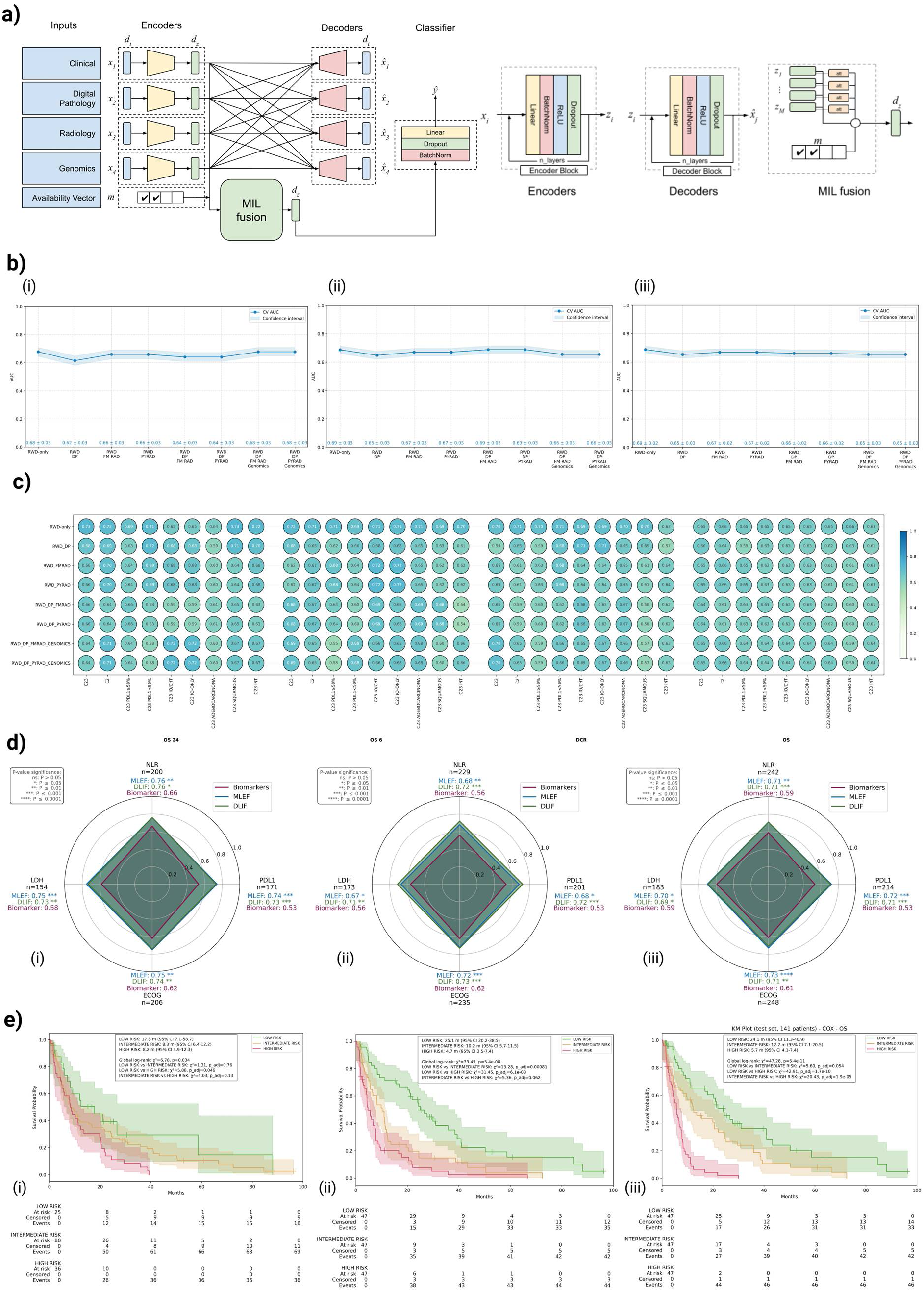
Deep Learning Intermediate Fusion (DLIF) performance and single-biomarker comparison. a) Schematic overview of the DLIF architecture designed to handle missing modalities; b) Classification performance of C23 unimodal, bimodal, and multimodal DLIF models for (i) OS24, (ii) OS6, and (iii) DCR on CV; c) Performance of DLIF classifier (AUC for OS24, OS6, DCR, and C-index for OS) across all subgroup analysis on the TEST, circle color indicates the model’s performance, while size corresponds to the size of pts cohort (equal across sub-analysis); d) Direct comparison between MLEF (RWD-only), DLIF (best performing multimodal model on CV) and single biomarkers for (i) OS24 (DLIF RWD FMrad), (ii) OS6 (DLIF RWD), and (iii) DCR (RWD DP FMrad) in the C23 cohort on TEST; performance assessed using AUC and tested with DeLong’s method, p-values were calculated using a two-sided t-test; e) Kaplan-Meier curves of low, intermediate and high risk patients; (i) separation by LIPI score on TEST subset with LIPI available (n=141), (ii) separation by tertiles from COX - MLEF on TEST LIPI subset (n=141), (iii) separation by terciles from COX - DLIF on TEST LIPI subset (n=141), p-values were calculated using the log-rank test. *OS-Overall Survival, DCR-Disease Control Rate, NLR - Neutrophil-to-Lymphocyte Ratio*

### MLEF and DLIF outperform single biomarkers

In the TEST for the combined C23 cohorts without missing biomarker data for OS24, MLEF RWD-only models and DLIF (best performing multimodal models on CV for each outcome) outperformed single biomarkers: PD-L1 (AUCs for MLEF vs DLIF vs single biomarker: 0.74 vs 0.73 vs 0.53), ECOG PS (0.75 vs 0.74 vs 0.62), LDH (0.75 vs 0.73 vs 0.58), and NLR (0.76 vs 0.76 vs 0.66), with *p*≤0.05 for all comparisons, **Fig. 3di**.

Similarly for OS6, both models again outperformed single biomarkers: PD-L1 (0.68 vs 0.72 vs 0.53), ECOG PS (0.72 vs 0.73 vs 0.62), LDH (0.67 vs 0.71 vs 0.56), and NLR (0.68 vs 0.72 vs 0.56), with *p*≤0.05 for all, **Fig. 3dii**.

In the TEST for DCR, MLEF RWD-only models outperformed single biomarkers: PD-L1 (0.72 vs 0.71 vs 0.53), ECOG PS (0.73 vs 0.71 vs 0.61), LDH (0.70 vs 0.69 vs 0.59), and NLR (0.71 vs 0.71 vs 0.59), with *p*≤0.05 for all, **Fig. 3diii**. The difference observed among AUC values for MLEF and DLIF models vs single biomarkers was not consistent across all other metrics (See ***Extended Data Fig.3**)*.

### Cox-MLEF and Cox-DLIF better stratify patients by risk groups than LIPI score

In the TEST subset with available LDH and NLR data (n=141), Cox-MLEF significantly separated Low vs High (OS: 25.1 vs 4.7 months, *p*=6.1e-08) and Low vs Intermediate risk groups (25.1 vs 10.2 months, *p*=0.00081); Intermediate vs High (10.2 vs 4.7 months) was not significant (*p*=0.062) **Fig. 3eii**. Cox-DLIF achieved significant separation for Low vs High (OS: 24.1 vs 5.7 months, *p*<0.0001) and Intermediate vs High (12.2 vs 5.7 months, *p*<0.0001), but not for Low vs Intermediate (24.1 vs 12.2 months, *p*=0.054), **Fig. 3eiii**. In contrast, LIPI only separated Low vs High risk (*p*=0.046) and failed in the other pairwise comparisons, **Fig. 3ei**. In the full TEST (n=273), Cox-MLEF analysis significantly distinguished all three risk groups (global *p*<0.000001), further highlighting the advantage of model-based stratification, ***Extended Data Fig. 3***. Overall, both Cox-MLEF (C-index 0.65, 95%IC 0.60-0.70) and Cox-DLIF (C-index 0.63, 95%IC 0.58-0.68) outperformed LIPI (C-index 0.55, 95%IC 0.50-0.60).

### Fairness evaluation of the MLEF RWD-only models revealed a high variability in sensitivity across centers

When using center as a protected attribute in TEST, the MH subgroup achieved significantly higher sensitivity for OS6 prediction (true positive rate (TPR) = 0.96) compared with other centers: INT (0.58), GHD, (0.62), VHIO (0.62), and SZMC (0.61), with *p*≤0.05, **Fig. 4ai**. For the DCR endpoint, a significant difference in sensitivity was also observed between MH (TPR = 0.96) and INT (TPR = 0.63) (*p*≤0.05, **Fig. 4ai**). In contrast, sensitivity was not statistically different when using sex as a protected attribute, **Fig. 4aii**. For false positive rate (FPR), no statistically significant differences were observed across centers or between sex (**Fig. 4bi-ii**). Fairness analysis in EXVAL showed that classification models performed equally across self-reported race and sex, ***Extended Data Fig. 4***. For OS survival model (Cox-MLEF), TEST C-index varied across centers (INT=0.69, GHD=0.61, VHIO=0.57, SZMC=0.67, MH=0.50); EXVAL C-index across self-reported race showed modest disparities when using cox (Black/African American=0.68, White=0.63, without statistical significance), ***Extended Data Fig. 4***.

### XAI analysis for MLEF RWD-only models aligns with established prognostic factors

Across endpoints, ECOG Performance Status (PS) greater than 0 and the presence of bone, liver, and brain metastases consistently emerged as strong negative prognostic factors for OS24 (**Fig. 4ci-ii**). NLR was additionally confirmed as important for OS24 and OS survival analysis, while high LDH influenced DCR non-responder predictions (***Extended Data Fig. 4***). Examples of local explainability graphs are presented for four patients from the TEST for OS24, representing True Positive (TP) (**Fig. 4ciii**), True Negative (TN) (**Fig. 4civ**), False Positive (FP) (**Fig. 4cv**), and False Negative (FN) (**Fig. 4cvi**). These graphs were further used for the clinical usability study (see **Fig. 5a**).

### Physician-XAI collaboration improved predictions

To estimate the benefit of the developed MLEF RWD-only baseline models (logistic regression (LR) and Cox) in supporting clinical decision-making, we conducted a clinical usability study involving 20 physicians (10 lung cancer experts and non-experts pairs). Each physician evaluated 10 real-world NSCLC cases from the TEST set, totaling 200 assessments. The detailed design is shown in **Fig. 5a**. Physicians were asked to estimate DCR and OS, split into five categories: <6, 6–12, 12–18, 18–24, ≥24 months, first without (Phase 1) and then with (Phase 2) support from the models and SHAP-based explainability (see *Online Methods* for more details).

For predicting DCR responders, Physician-XAI collaboration improved sensitivity for all physicians from 0.72 (95% CI: 0.64–0.80) to 0.87 (95% CI: 0.79-0.92), *p*=0.0011, accuracy from 0.57 to 0.65, *p*=0.0431, and F1-score from 0.67 to 0.75, at the expense of slightly lower specificity (**Fig. 5bi**). Both non-experts and experts benefited from the XAI use (sensitivity from 0.77 to 0.90, *p*=0.0209, **Fig. 5biii** for non-experts and from 0.68 to 0.83, *p*=0.0201, **Fig. 5bii** for experts), with similar trends in accuracy and F1-scores. Overall, a 37% increase in the likelihood of correct DCR prediction for all physicians was observed when using XAI, though not statistically significant (*p*=0.1250). Similarly, a non-statistically significant improvement of +53% and +23% correct DCR predictions for non-experts and experts was observed, respectively. The agreement between experts and non-experts improved from slight (κ=0.11) to moderate (κ=0.48) from Phase 1 to Phase 2. Overall, physicians frequently adopted correct XAI suggestions (74.5%), but experts were more likely to follow incorrect ones (72.2% vs 63.6% for experts and non-experts, respectively) **Fig. 5c**.

In the OS estimation task, the use of XAI increased the probability of correctly predicting OS by 36% for all physicians, 14% for experts and 61% for non-experts. In addition, we developed a scoring system (see *Online Methods* for details) and the total score for correctness for all physicians increased by 4.5%, for non-experts by 6.5% and for experts by 2.5%, **Fig. 5di**. In Phase 1, 16.5% of the physicians’ evaluations were concordant with the true OS; such proportion increased to 22.5% after using the XAI (**Fig. 5dii**). Finally, the inter-physician agreement improved from slight to fair (κ from 0.14 to 0.34), see ***Supplementary Information 2*** for full analysis.

## Discussion

I3LUNG is the largest international, real-world multimodal study in NSCLC IO and AI, encompassing 2396 patients from six centers in six countries, including 339 patients with complete data modalities. By combining RWD, CT scans, DP, and genomics, together with fairness audits, XAI and clinical usability, the I3LUNG PDSS offers a uniquely comprehensive dataset and framework for the development and validation of immuno-oncology multimodal predictive models.

Our analysis demonstrated that the AI-based models with routinely available RWD-only outperformed single biomarkers (PD-L1, ECOG PS, NLR, LDH) and the composite LIPI score used in clinical practice. In the combined C23 cohort and across end-points, MLEF models achieved consistent performance (CV AUC up to 0.77), with superior results (CV AUC up to 0.88) for first-line IO in predicting long-term survivors (OS24). A lower performance was observed with Cox-MLEF, yielding a C-index up to 0.74 (C2). These results align with prior large-scale, RWD-based efforts such as SCORPIO^16^, which achieved AUC values of 0.76 (OS) and 0.64–0.71 (clinical benefit) using RWD from >9,000 patients to outperform traditional biomarkers (TMB, PD-L1). Our unimodal models based on FMRAD, DP, or GEN alone performed similar to other single modality efforts in literature. For instance, Saad *et al.*^10^ reported a C-index of 0.75 using DEEP-CT, a deep learning model developed with 976 baseline chest CT scans, combined with clinical data. Rakaee *et al.*^11^ achieved an AUC of 0.75 internally and 0.66 in external validation with H&E slides-based prediction (n=958). A great effort to overcome these challenges is represented by the LORIS model developed by Chang *et al.*^17^, a pan-cancer logistic regression framework integrating six clinical, TMB and pathologic features. LORIS demonstrated strong performance (AUCs up to 0.83 for IO response), and was validated on multiple unseen datasets, achieving an AUC of 0.74 for the NSCLC specific-model. However, LORIS relies on TMB, which is rarely available in routine clinical practice, and does not leverage any imaging data. Collectively, these studies suggest that, while promising, uni- and bi-modal frameworks still fail to capture the full complexity of IO response, reinforcing the rationale for multimodal integration.

In our analysis, multimodal integration did not consistently outperform unimodal RWD. This may reflect both the heterogeneous quality of data across centers and the strong predictive signal already embedded in structured clinical features. Real-world genomic data did not provide any added value to model performance, likely due to feature encoding challenges and sparse data availability. Overall, DLIF and MLEF performed comparably, reinforcing the value of efficient, interpretable, lower-complexity models. Adding modalities did not improve DLIF performance, reflecting its vulnerability to missing data. In contrast, MLEF performed better in homogeneous subgroups with complete multimodal data, the most effective combination being RWD+FMRAD+DP. However, the incremental value of multimodality was not consistent. Specifically, multimodality did not enhance performance in subanalyses where RWD already provided strong predictive signals, highlighting that its added value lies primarily in settings where RWD underperforms. Performance of both MLEF and DLIF remained stable in TEST, but declined in EXVAL, likely reflecting population shifts (e.g., higher long-term survival in UOC) and underscoring generalizability challenges. Previous multimodal studies have faced similar constraints, including small cohorts, single-center designs, or lack of external validation. For instance, Vanguri *et al.*^13^ developed DyAM, integrating CT radiomics, PD-L1 IHC, and genomics to predict IO response in NSCLC, achieving AUC of 0.80 (95% CI: 0.74–0.86). While representing a pioneering effort in developing a rigorous methodology for handling missing data modalities, the study was limited by its modest sample size (n=247; 81 fully multimodal), single-center design, and absence of TEST or EXVAL. More recently, Captier *et al.*^14^ combined PET/CT, pathology, transcriptomics, and clinical data from 317 NSCLC patients (80 fully multimodal), achieving an AUC 0.81 and C-index 0.75 with late-fusion models. In this case, the lack of external validation limits generalizability, and transcriptomics remains a non-standard, costly modality for standard clinical workflows. Of note, the models published in these studies cannot be directly applied to our work, as they often rely on data not routinely available across diverse healthcare systems (e.g., TMB^13,17^), or because access to the models and used data requires a data sharing agreement.^10,11,16^ Beyond predicting IO^13,17^ or IO/CHT^10,14^ efficacy, emerging work by Saad *et al*.^18^ extends these efforts towards treatment selection. Their ML scoring systems combining genomic and clinical features identified patients more likely to benefit from the addition of CHT to IO. This shift from prediction to treatment optimization underscores that AI model evaluation should address not only model performance, but also clinical utility.

In our work, fairness audits across protected attributes revealed variability in sensitivity across centers, likely driven by heterogeneous data quality and TEST sample imbalances (detailed in ***Extended Data Fig. 1***). While differences in Cox-MLEF performance by self-reported race were not statistically significant, they underscore the need for fairness-aware calibration and site-specific validation before clinical deployment. XAI findings aligned with established prognostic factors, reinforcing the clinical plausibility of the RWD-only MLEF models.

The clinical usability study demonstrated that XAI applied to RWD-only models can meaningfully support physician decision-making. Sensitivity for DCR prediction improved from 0.72 to 0.87 with XAI, with parallel gains in accuracy and F1-score. This sensitivity gain is clinically valuable, reducing missed opportunities for patients likely to benefit from IO. Non-experts achieved greater relative improvements than experts, demonstrating the potential of XAI to assist less experienced or non-specialist clinicians. OS prediction also improved modestly in Phase 2, with an overall gain of 36% among all physicians. Such improvement may be especially impactful in community settings lacking thoracic oncology expertise. Although retrospective, the clinical usability study provides a reproducible and structured reference framework for future evaluation of PDSS usefulness and XAI analysis in real-world clinical settings. The analysis of questionnaires collected during the study will be published separately and will inform refinement of the XAI approaches, supporting the development of an interface suitable for real-world, prospective clinical use.

This study has limitations. First, these first analyses relied on retrospective data from patients treated with first- and later-line IO. Furthermore, the heterogeneity and incompleteness of the available modalities, reflecting real-world clinical variability, likely contributed to reduced model performance. Concerning genomics, a limitation lies in the composite “driver” feature that was used to group multiple actionable alterations (*EGFR*, *ALK*, *RET*, *ROS1*) into a single binary variable, to address data sparsity. While mitigating missingness, this approach may have obscured the distinct effects of individual alterations on IO response and survival. More comprehensive and standardized genomic datasets will be required to fully explore their potential. Another limitation is that radiomic features were extracted only from primary lesions, potentially overlooking valuable information from metastatic sites. Because multimodal MLEF models outperforms RWD-only just in specific subpopulations, we decided to focus the rest of the analysis, post-hoc XAI and fairness, on RWD-only MLEF models. Finally, we have not yet explored the use of large-scale, generalizable vision-transformers for multimodal data integration. However, as recent research efforts are shifting from single-task models to foundation and generative AI approaches, our next steps will include testing vision-transformers to improve multimodal data integration and automation. One notable example is MSK-CHORD by Jee *et al.*^19^, which leveraged transformer-based NLP on a >24,000-patient clinico-genomic dataset (7.800 NSCLC), reaching C-indexes of 0.58-0.83 across cancers. Similarly, Xiang *et al.*^20^ developed MUSK, a vision-language model trained on 11,000 patient tokens, predicting IO response in NSCLC with AUC 0.77 (n=118), although limited to a single cohort and pathology combined with clinical data. These approaches, while promising, require large datasets, and XAI remains a major challenge. Despite these limitations, our study represents an early prototype towards the development of a global platform for NSCLC multimodal data integration into clinical practice.

In conclusion, the first I3LUNG PDSS demonstrates that single biomarkers, composite scores, or subgroup analyses derived from randomised clinical trials are no longer sufficient to capture the complexity of IO response. AI-based integration, even when limited to RWD, provides superior predictive power, with multimodal models adding clear value in defined subgroups where traditional markers fall short. By leveraging data from six international centers encompassing diverse healthcare systems and imaging protocols, this study provides a framework that mitigates overfitting, evaluates model generalizability and fairness, and assesses clinical usability. These efforts address key challenges inherent to retrospective, heterogeneous real-world evidence with missing modalities and align with the FUTURE-AI^21^ framework, advocating for human-in-the-loop design.

Together, these findings establish I3LUNG as a large-scale benchmark for applying ML and DL methods, including foundation models in digital pathology and radiology, to real-world immuno-oncology. Future work will extend validation to the ongoing prospective multimodal (>2,000 patients) and multi-omics (>200, 11 omics) cohorts, explore multimodal vision-transformers, and refine usability tools to bridge expertise gaps and support seamless clinical integration.

## Online Methods

The I3LUNG project (NCT05537922) employs a two-phase approach to develop a medical device aimed at predicting immunotherapy efficacy in NSCLC patients. This framework integrates multimodal data, including real-world clinical data and multi-omics datasets, through retrospective and prospective study phases. In this manuscript we report the results from the retrospective study phase, which is focused on extracting preliminary knowledge to construct a retrospective predictive model for IO outcomes. This model served as the foundation for developing the initial version of the PDSS.

### Patient enrollment

This study included retrospective data from 2396 patients with stage IIIC-IVB NSCLC who received IO-based therapy between September 2012 and October 2023 across six international institutions: Istituto Nazionale dei Tumori (INT), Italy; LungenClinic Grosshansdorf (GHD), Germany; Metropolitan Hospital (MH), Greece; Shaare Zedek Medical Center (SZMC), Israel; Vall d’Hebron Institute of Oncology (VHIO), Spain; and the University of Chicago (UOC), USA. Patients with available baseline data and known clinical outcomes (response to IO according to RECIST, OS) were included in the analysis. All patients were enrolled as part of the I3LUNG international, multicenter, retrospective and prospective observational study (NCT05537922). Eligibility criteria included adults aged 18 years or older, and a histologically confirmed diagnosis of stage IIIC-IVB NSCLC according to the 8th edition of the TNM staging system. The study was conducted in compliance with the Declaration of Helsinki and Good Clinical Practice (GCP) guidelines. Ethical approval was obtained from an independent ethics committee at each participating site.

### Clinical data

The study population was subdivided into three distinct patient cohorts based on NSCLC disease stage:

● *Cohort 1* – Curative Setting: Includes patients with stage III NSCLC treated with curative-intent chemoradiation therapy (concomitant or sequential) followed by maintenance IO.
● *Cohort 2* – Non-Curative Setting, First Metastatic Line: Includes patients with advanced or metastatic NSCLC who received first-line IO, either alone or in combination with CHT or other therapeutic agents.
● *Cohort 3* – Non-Curative Setting, Later Lines: Includes patients with advanced or metastatic NSCLC treated with IO in any line beyond first-line, either alone or in combination with CHT or other agents.

For patients who received IO in both curative and non-curative settings, their data were duplicated and included in both Cohort 1 and either Cohort 2 or Cohort 3, depending on their metastatic treatment line. Consort flow is shown in ***Extended Data Fig. 1ai***.

The first step in the data curation process involved hypothesis-driven feature selection. Initially, we extracted more than 11000 features from the electronic Case Report Forms (eCRF) from the centralised I3LUNG platform. These features were then categorised into two main groups: (i) Baseline Features: RWD available before the administration of the first IO cycle; (ii) Treatment-Related Features: Data primarily related to treatment efficacy and toxicity. Due to the high dimensionality of features, a blinded review process was conducted by two independent groups of oncologists (LP and CS vs AS and CG) to refine the feature selection. Each group consisted of two oncologists, including one with lung cancer expertise. A total of 227 features were selected in common by both groups. The selected baseline IO features were further divided into: (i) Descriptive Features – Used for exploratory analysis and sub-model development, and (ii) Predictive Features – Used to train the models. The final hypothesis-driven feature selection from predictive features was performed by MG and AP, a senior oncologist with extensive experience in lung cancer treatment, who refined the selection by reviewing, summarizing, and consolidating previous choices, resulting in a final set of 9 RWD. The second step focused on data cleaning and quality control, with a primary emphasis on: (i) cleaning free-text inputs to ensure consistency, and (ii) handling inconsistencies in dates and numerical values. As part of the quality control process, we conducted three iterative rounds of validation with the participating centers, requesting corrections and clarifications for identified data inconsistencies. This iterative approach ensured that missing values, inaccurate entries, and discrepancies across datasets were resolved as much as possible, improving overall data reliability and consistency.

### Clinical end-points

We used two types of clinical endpoints to evaluate treatment efficacy: response outcomes and survival outcomes. Response outcomes were assessed using the RECIST v1.1, leading to the definition of key response metric - Disease Control Rate (DCR). Patients were classified as:

a. Non-responders (Class 0): Progressive Disease (PD)
b. Responders (Class 1): Stable Disease (SD), Partial Response (PR), and Complete Response (CR)

We selected Overall Survival (OS), the time from initiation of IO to death or last follow-up to assess treatment durability.

The endpoints of the study were the identification of long responders - Overall Survival at 24 months (OS24), poor responders - Overall Survival at 6 months (OS6), and DCR for classification tasks, and the OS for survival analysis. Full statistical analysis of the RWD across centers and cohorts is shown in ***Supplementary Information 3***.

### CT scans

CT scans were collected in DICOM format and underwent quality control, harmonization and finally, segmentation. Given the multicentric nature of the study, CTs acquisition was heterogeneous. To ensure effectiveness of downstream analysis, a number of exclusion criteria has been applied, such as CTs with missing metadata, usage of non standard or soft tissue kernels, slice thickness out 1-5mm, clinical exclusion (e.g. patients with concurrent malignancies), corrupted DICOM, consort flow for CT scans is shown in ***Extended data Fig.1aii***. When available, contrast–enhanced scans were preferred. Segmentations were performed using 3D slicer, relying on a hybrid approach combining semi-automated and manual refinement techniques. The segmentation was done by a single trained radiologist. Problematic cases, difficult segmentations were reviewed by a senior radiologist (BN) who reviewed and approved the segmentation. Thus, the segmentation process was uniform for all cases with no inter-operator variability. The final segmentation masks were exported as NRRD files by the team at SZMC. Available metadata analysis, with focus on manufacturer, slice thickness and contrast information of the included CT scans (by center) is shown in ***Supplementary Figure 2ai-iii***. To assess the robustness and discriminative power of radiomic features, we applied a contour randomisation approach to each lesion segmentation, which the radiologist team at SZMC had manually delineated. For each lesion, we generated 10 randomised versions of the original mask, resulting in 11 segmentations per lesion (1 original + 10 perturbed). Radiomic features were extracted from each of these segmentations using PyRadiomics, resulting in 1,400 features per setting. To evaluate feature stability and relevance, we applied a statistical framework based on the inverse coefficient of selection *IS_f_*. This metric captures both intra-sample robustness σ^2^_*inter,f*_ and inter-sample discriminability σ^2^_*inter,f*_. Let *l* ∈ {1, . . ., *L*}, *p* ∈ {1, . . ., *P*}, *f* ∈ {1, . . ., *F*} denote lesion, perturbation, and feature indices, respectively. Formally, the inverse coefficient of selection *IS_f_* is defined as follows:

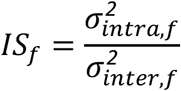

where

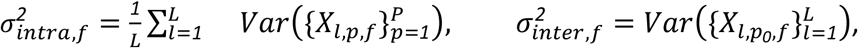

and *X_l,p,f_* is the value of the feature f for lesion l, and perturbation p. σ*_intra_* variance was computed by assessing the average variability of each feature across all perturbation settings for a given lesion, where lower intra-variance indicates higher robustness. σ_*inter*_ variance was computed as the variance of each feature across all patients in the original setting, serving as a measure of discriminative power, where higher indicates better. Finally, *IS_f_* is defined as the ratio of intra- and inter-variance, where lower values indicate more stable and discriminative features. Features were ranked by *IS_f_*, and we then applied an iterative selection strategy to avoid redundancy: at each step, the next best-ranked feature was added only if it was not highly correlated with any of the already selected features. This process ensured the selection of a compact, non-redundant set of 128 robust and informative radiomic features.^22^ Example of expert CT segmentation with perturbations is shown in ***Supplementary Figure 2b***.

Regarding the FMRAD approach, to extract high-level imaging features from CT scans, we used a domain-specific FM developed for cancer imaging biomarker discovery.^23^ This model was pre-trained using contrastive learning on a large dataset of 11,467 radiographic lesions, enabling it to learn generalized and transferable representations of tumor morphology. For our study, we applied this FM to volumetric image patches of size 50 × 50 × 50 mm, extracted around the centroid of the segmented primary lesion. The lesion center was determined from the segmentation mask, and the surrounding cubic volume was resampled and standardized as required by the FM input specifications. The resulting embeddings served as compact and expressive imaging descriptors for subsequent multimodal integration and predictive modelling.

### Digital Pathology

We compiled a total of 998 histopathological slides for our study, sourced from five institutions: 117 slides from GHD, 364 from INT, 81 from MH, 147 from SZMC, and 191 from UOC. Of these, 64 slides were excluded from further analysis due to quality control concerns, such as insufficient tissue, physical damage (i.e., scratched slides), poor staining quality, and other technical artifacts that could compromise downstream analysis (Consort flow in ***Extended data Fig1aiii***). Each remaining slide was individually reviewed, with systematic annotation of pen markings, scanner-induced artifacts, and out-of-focus regions. For tumor region identification, we trained a segmentation model using the TCGA dataset. Tile extraction was performed at 20x magnification, followed by Reinhard normalization, targeting only areas within the segmented tumor regions, while avoiding the previously annotated artifacts. We used Prov-GigaPath^24^, a slide-based pretrained foundation model: first, to encode each extracted tile into a vector; then, to aggregate each bag of vectors into a single embedding representative of the Whole Slide Image.

### Genomics data

To include genomics data into our models, we first identified a core set of primary driver genes relevant to NSCLC: *KRAS*, *TP53*, *STK11*, *ALK*, *EGFR*, *RET*, and *ROS1.*^25^ Pathogenic alterations were annotated using three widely recognized databases: ClinVar^26^ (via Varsome), Cancer Hotspot^27^, and OncoKB^28^. Given minor inconsistencies among databases, an alteration was classified as pathogenic only if at least two out of the three sources agreed. For model construction, we encoded individual binary features for *KRAS*, *TP53*, and *STK11*, where 1 indicated a detected mutation, 0 indicated a wild-type allele, and NA indicated missing testing information. Additionally, we created a composite “driver” feature to capture actionable alterations (*EGFR*, *ALK*, *RET*, or *ROS1*), assigning a value of 1 if any of these were altered upon testing, 0 if at least one was tested and not altered, and NA if no testing data were available. This approach helped reduce missingness, given the common practice of testing these genes together in clinical workflows, and aimed to preserve signal strength for modelling of IO response and survival.

### Development of the AI tool

We evaluated two main AI approaches for treatment outcome prediction: Machine Learning Early Fusion (MLEF) and Deep Learning Intermediate Fusion (DLIF). The data preprocessing and analysis pipelines were identical for both methods.

As part of the RWD used for model training, we included a curated set of clinical and biological features selected by two expert lung oncologists (MG and AP) for the main analysis reported in the manuscript. These included: patient characteristics (sex, ECOG PS, smoking status), molecular features (PD-L1 expression), metastatic sites (bone, liver, brain), and laboratory values (NLR, LDH). Details on feature encoding are provided in ***Supplementary Table 2***. In addition, we have run the data-driven approach by selecting RWD using LASSO starting from 227, since there was no significant difference in results (see ***Supplementary Table 3)*** here we report results obtained by using 9 selected features. Missing values were handled using multivariate imputation via the IterativeImputer (1.6.1). We used CV with each fold corresponding to a different center (i.e., Leave-One-Centre-Out (LOCO)), resulting in a total of 5 CV folds.

We performed the main analysis on combined Cohort 2 and Cohort 3 to increase the cohort size. Additionally, we performed several subgroup analyses to evaluate performance across clinically homogeneous subpopulations. These included models trained on: cohort 2, IO-only, IO/CHT, PD-L1≥50%, PD-L1<50%, squamous histology, adenocarcinoma histology, and site-specific cohorts; summary of all analyses is in ***Supplementary Table 1*.**

For benchmarking, we compared model performance against individual clinical biomarkers, including PD-L1 expression, NLR, LDH, and ECOG PS.

#### Machine Learning Early Fusion (MLEF)

Before training the models, we performed feature selection using LASSO regression for FMRAD due to the high embedding dimensionality (more than 4000 in length). For other modalities we started from the initial number of features (as shown in **Fig. 1a**), we concatenated them across all modalities and applied LASSO to select the most relevant predictors.

We implemented and evaluated two ML classifiers: (i) Logistic Regression (LR), and (ii) Random Forest. We selected the optimal hyperparameters by maximizing the AUC during model tuning. The best MLEF classifier was selected using CV AUC. All performance results include 95% CI, and comparisons across models were conducted using DeLong’s test for AUC differences, with p-values calculated using two-sided t-test, n.s. - not significant, * p≤0.05, ** p≤0.01, *** p≤0.001, **** p≤0.0001. To evaluate model performance against single biomarkers, we focused on a subset of TEST for which biomarker measurements were available.

For the survival analysis, we implemented a Cox Proportional Hazards (Cox) model as baseline, using the lifelines package in Python^29^. Additionally, we evaluated two ML-based survival models (Random Survival Forest (RSF), Gradient Boosting Survival (GBS)) implemented with scikit-survival^30^. Since no difference between models were noted, all the results for survival analysis refer to the Cox model.

Because the MLEF pipeline cannot handle patients with missing modalities, all multimodal models exclude these patients. For transparency, each MLEF multimodal model was reported along with the number of patients included in the analysis, as well as the best model for that analysis, and it was compared to the unimodal RWD-only matched model using the same patient subset.

#### Deep Learning Intermediate Fusion (DLIF)

Our second approach, DLIF, was designed to address the common issue of missing data modalities across patient cohorts. We implemented a supervised intermediate fusion pipeline capable of handling both classification and time-to-event survival prediction tasks. This architecture integrates RWD, DP, RAD and G using an Attention-Based Multiple Instance Learning (MIL) framework^31^. In this framework, each patient is represented as a bag of vectors, allowing a variable number of instances per patient. This design is particularly well suited to complex oncological datasets, such as the I3LUNG retrospective cohort (**Fig. 1b-d**), where full multimodal coverage was available only for a subset of patients. To meet the MIL architecture’s requirement for fixed-size input vectors, we implemented modality-specific encoders that project each modality into a shared embedding space. Beyond aligning dimensionality, it was essential to ensure that these embeddings occupy the latent space in a consistent and interpretable manner. To achieve this, we included a cross-modal reconstruction loss during training, encouraging the model to reconstruct one modality from another. Both loss functions combine a primary task-specific loss with a reconstruction loss component:

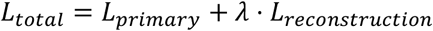

where λ is the reconstruction weight parameter. The reconstruction loss encourages the model to learn meaningful cross-modal representations by requiring it to reconstruct missing modalities from available ones. For a batch with *N* samples and *M* modalities, let: *x*^(*i*)^_*m*_ be the input feature for sample (*i*) and modality *m*, *x̂*^(*i*)^_*m,j*_ be the reconstruction of modality *j* from modality *m* for sample *i*, *M*^(*i*)^ ∈ {*0*,*1*}^*M*^ be the modality mask indicating available modalities for sample *i*, the reconstruction loss is computed as:

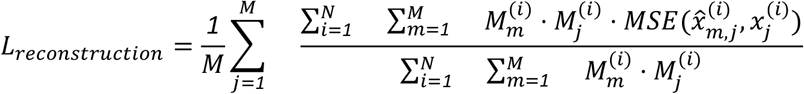

where the double summation ensures only valid reconstructions (where both source and target modalities are available) contribute to the loss, the normalization by the count of valid reconstructions prevents bias when modalities are missing and MSE is computed as: 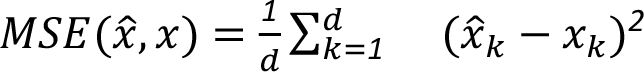 where d is the feature dimension. For multimodal classification tasks, the primary loss is cross-entropy:

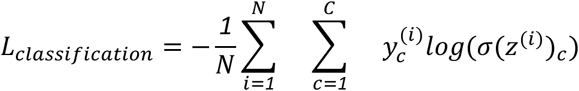

where *C* is the number of classes, *y*^(*i*)^_*c*_ is the one-hot encoded ground truth for sample *i* and class *c*, σ(*z*^(*i*)^) is the softmax of the model’s logits for sample *i*, optional class weights *w* ∈ *R^C^* can be applied:

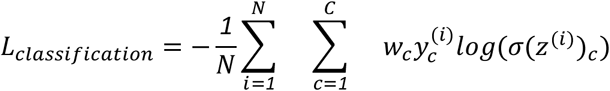

For survival analysis, the primary loss uses the Cox Proportional Hazards model:

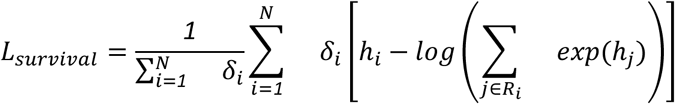

where *h_i_* is the log-hazard prediction for sample *i*, δ_*i*_ is the event indicator (1 if event occurred, 0 if censored), *R_i_* = {*j: T_j_* ≥ *T_i_*} is the risk set (samples with survival time ≥ *T_i_*), samples are sorted by descending survival time for computational efficiency. To prevent overflow in the exponential operations, the log-sum-exp trick is used:

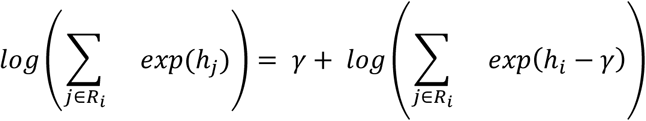

where γ = *max_j∈R_i__h_j_*.

This design constraint promotes information alignment across modalities and effective fusion through the MIL attention mechanism.

### Fairness evaluation

To assess fairness, we conducted a post-hoc auditing focusing on potential biases across subgroups in TEST and EXVAL. We used center and sex as protected attributes for TEST and race and sex for EXVAL.

To quantify fairness, for classification models, we used equalized odds, which requires that both true positive rate (TPR, also known as sensitivity) and false positive rate (FPR) are consistent across subgroups. TPR and FPR were calculated for the best performing RWD-only MLEF model.

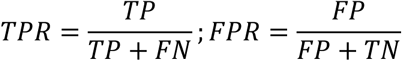

For survival analysis models, we applied Parity in C-index, to verify whether model discrimination remained consistent across centers.

Fairness was evaluated using statistical tests to compare model performance across subgroups. For the classification tasks we performed a permutation test with 1000 iterations, for the survival task we used bootstrapping with n=1000. In both cases p-values were calculated using a two-sided test for comparison between the two groups.

### Explainability

Explainability analysis was conducted using SHAP^32^. For each clinical endpoint, selecting the best performing model, we generated summary SHAP plots trained on the TRAIN and applied both on the TRAIN and TEST. In addition, we produced individual waterfall plots for each patient in the TEST. These graphs are included either in the main text (**Fig. 4c**) or ***Extended Data Fig. 4***, and were also used for the clinical usability study.

**Figure 4.**
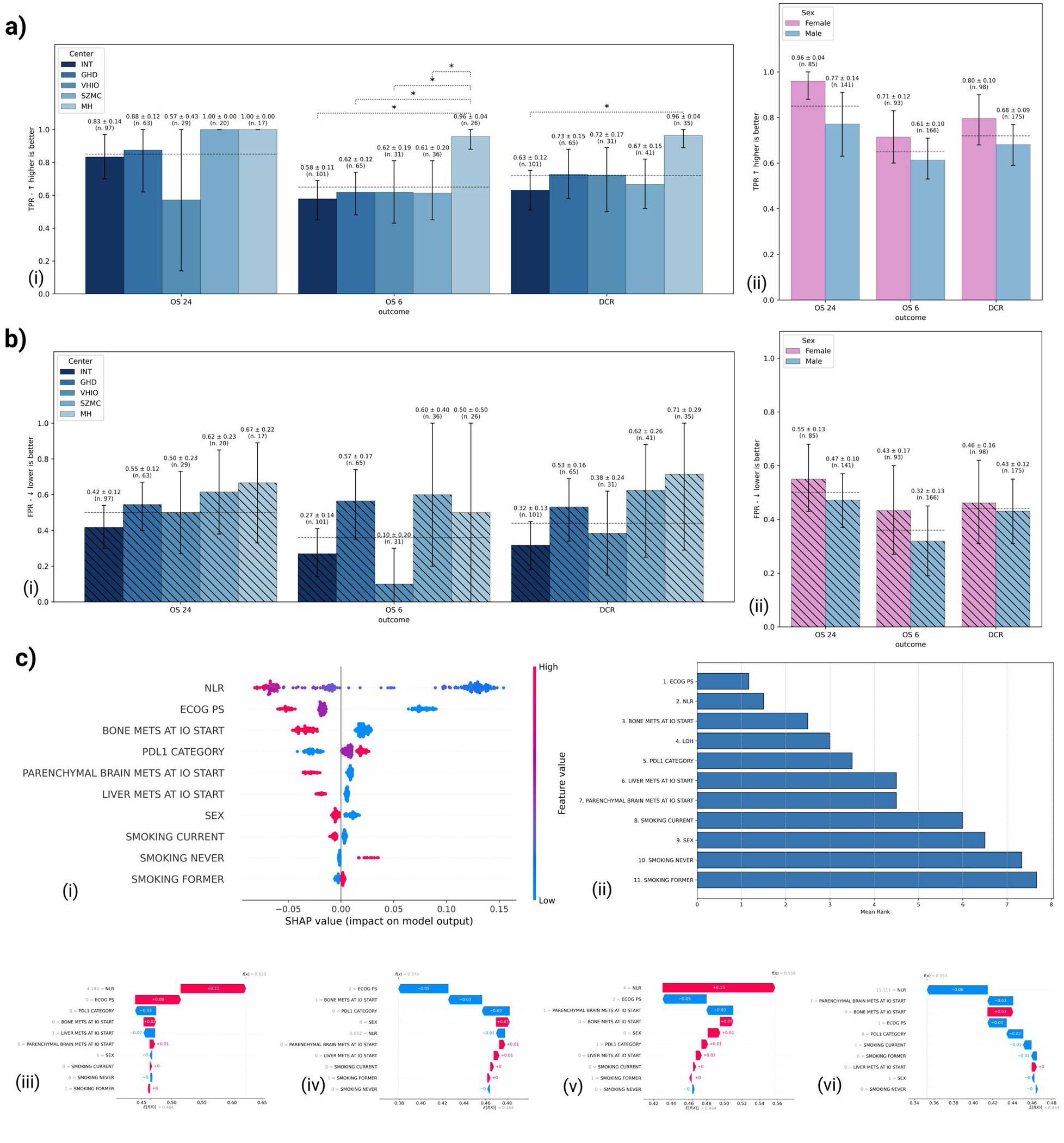
Fairness (equalised odds) and XAI analysis of RWD-only MLEF models. a) Sensitivity (TPR) in TEST across protected attributes, (i) center and (ii) sex, black dashed line indicates TPR value across full TEST set, p-values were calculated using a permutation test with 1000 iterations and a two-sided test for comparison between the two groups, n.s. - not significant, * p≤0.05, ** p≤0.01, *** p≤0.001, **** p≤0.0001; FPR in TEST across protected attributes, (i) center and (ii) sex, black dashed line indicates FPR value across full TEST set, p-values were calculated using a permutation test with 1000 iterations and a two-sided test for comparison between the two groups, n.s. - not significant, * p≤0.05, ** p≤0.01, *** p≤0.001, **** p≤0.0001; b) Post-hoc global explainability for OS24 model, (i) global summary SHAP plot for TEST; (ii) mean-ranked feature importance based on SHAP global values across OS24, OS6, and DCR outcomes; local SHAP explanation for four patients from TEST, True Positive (iii), True Negative (iv), False Positive (v) and False Negative (vi). *TPR-True Positive Rate, FPR-False Positive Rate*

To identify the most influential features across outcomes, we created a summary graph (**Fig. 4**) by calculating mean-ranked feature importance based on SHAP global values across all outcomes, the higher the rank the higher the feature importance across analysis.

### Clinical usability

The primary objective of the clinical usability study was to assess the I3LUNG tool’s clinical applicability. The clinical usability patient cohort consisted of 100 patients, of whom 35 had CT scans available, while 42 had H&E slides available. Patients were stratified into subgroups randomly.

The study involved 20 medical oncologists, consisting of 10 expert lung oncologists and 10 non-expert lung oncologists from the 5 centers (INT, VHIO, GHD, MH and UOC) from the consortium. Each patient was evaluated by one lung expert and one non-lung expert oncologist, with each physician evaluating a total of 10 patients.

The study design consisted of 2 evaluation phases (**Fig. 5**). In phase 1, medical oncologists had access to important clinical data, when available segmented CT scan slice, and DP slide. In phase 2, they had access to the available data and models output with global and local SHAP explanations. Supporting material provided to medical oncologists who were participating in the study is available in the ***Supplementary Information 4***, including the tutorials on how to read the model’s explanation (tutorials for global and local explainability with SHAP). Patient evaluations were conducted based on two key outcomes: prediction of treatment response (DCR) and of survival (OS).

**Figure 5:**
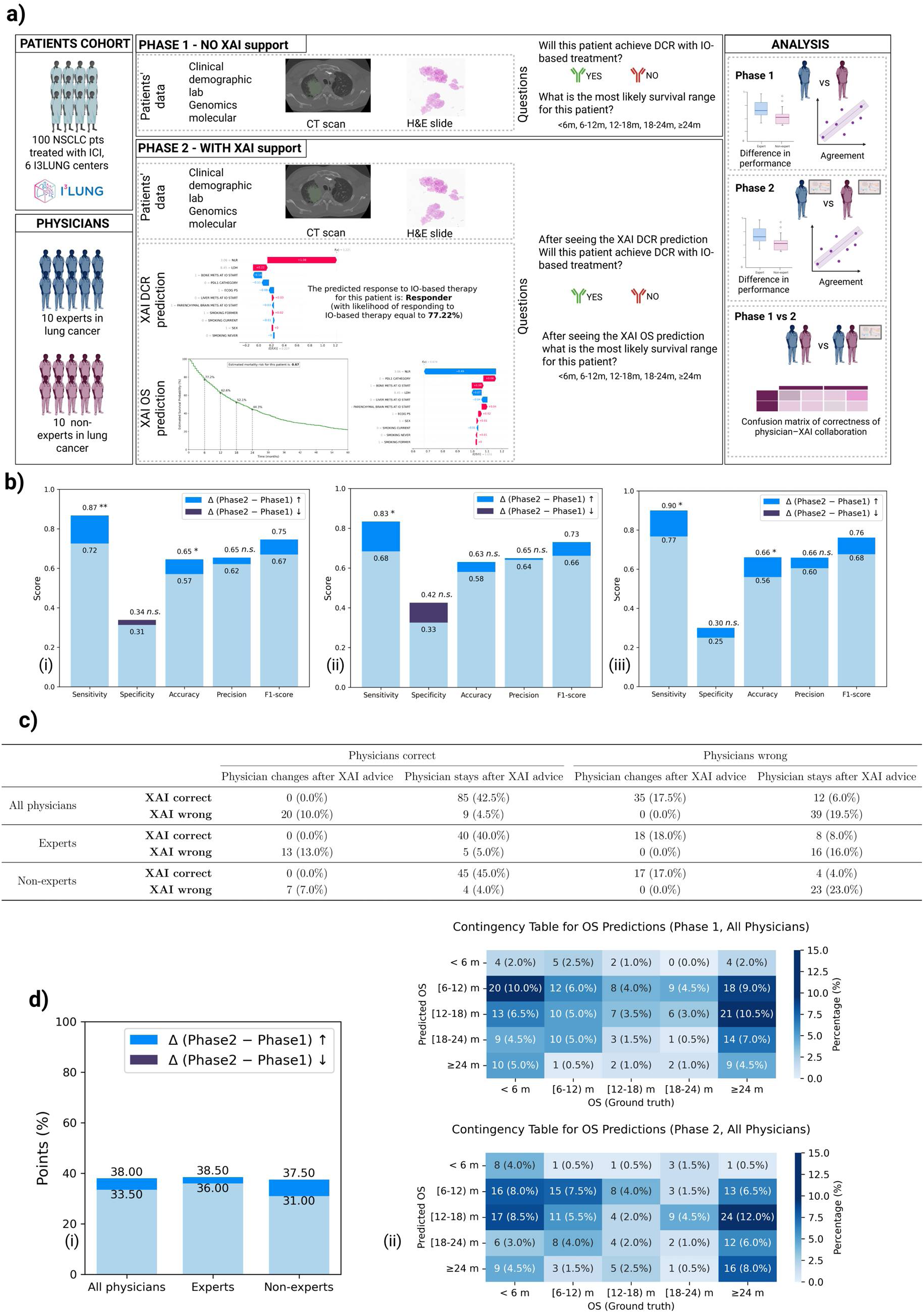
Clinical usability of MLEF RWD-only models. a) Schematic overview of clinical usability study. We selected 100 patients (80 from TEST and 20 from EXVAL set), 10 lung cancer experts and 10 non-experts evaluated 10 patients each for two endpoints DCR and OS, first in Phase 1 using just clinical data and in Phase 2 using clinical data and RWD-only MLEF (LR and Cox) output together with global and local explainability for each patient. We have analysed the difference in performance in Phase 1 and 2 among experts and non-experts, their agreement and, finally, the difference in performance in Phase 1 vs 2; b) Metrics summary for Phase 1 vs Phase 2 for DCR prediction for: (i) all physicians, (ii) experts, and (iii) non-experts, with p-values calculated using McNemar’s test, n.s. - not significant, * p≤0.05, ** p≤0.01, *** p≤0.001, **** p≤0.0001, statistical test for F1 was not performed; c) Confusion matrix of correctness of human-XAI collaboration in Phase 2 for DCR, illustrating whether physicians changed or maintained their prediction after XAI advice; d) Comparison of Phase 1 vs 2 for predicting OS range (i) Total score for OS predictions for Phase 1 and 2 according to physicians’ expertise, (ii) Contingency table for OS predictions in Phase 1 (top) and 2 (bottom) for all physicians. *XAI - eXplainable AI, OS-Overall Survival, DCR-Disease Control Rate*

For treatment response, physicians categorised each patient as either a responder (CR, PR, or SD) or a non-responder (PD), following DCR RECIST criteria^33^. For survival, physicians assigned each patient to one of five predefined OS time intervals: <6, [6–12), [12–18), [18–24), and ≥24 months whereas the tool generated an individual OS probability curve for each patient.

To evaluate the impact of XAI and physician expertise on prediction performance, we performed a series of univariable logistic regression analyses. For DCR two models were run and in both cases the outcome was binary, indicating prediction success (1: correct prediction; 0: incorrect). In the first analysis to assess the effect of XAI, we used phase as the covariate (0: no XAI, Phase 1; 1: with XAI, Phase 2), for the second analysis to assess the effect of physician expertise, we used expertise level as the covariate (0: non-expert; 1: expert). For OS prediction, success was defined as whether the predicted OS range overlapped with the true OS ±25% interval (1: overlap; 0: no overlap). Similar two univariable logistic regression models were applied first to test the effect of XAI use, where phase was used as the covariate, second to assess the role of expertise the covariate was (0: non-expert; 1: expert). All models were run for the full physician group, and separately for experts and non-experts. All the results were presented as Odds Ratio (OR) and AUC with their 95% Confidence Interval (95% CI) and are shown in the ***Supplementary Information 2***.

The degree of agreement between the expert and non-expert physicians’ DCR and OS evaluations were assessed by computing the Cohen’s Kappa and the weighted Cohen’s Kappa, respectively. Based on the observed Kappa, the degree of concordance were considered as follows:

- 0: no agreement

- [0.01 - 0.20]: slight agreement,

- [0.21 - 0.40]: fair agreement,

- [0.41 - 0.60]: moderate agreement,

- [0.61 - 0.80]: substantial agreement,

- [0.81 - 0.99]: near perfect agreement,

- 1: perfect agreement.

McNemar’s test for paired data was employed to assess any differences between experts and non-experts in the discordant predictions in phase 1 and phase 2.

Furthermore, metrics such as sensitivity, specificity, accuracy and precision were computed for the DCR prediction according to phase and physicians’ expertise. The 95% CIs of these metrics were calculated by using the Clopper-Pearson’s exact binomial method. McNemar’s test for paired data was employed to assess any statistically significant differences. The F1-score was derived according to phase and physicians’ expertise for the DCR prediction. Its 95% CI was computed by using bootstrap methods with 1000 replications.

Due to the time-to-event nature of the OS endpoint (i.e., it is not possible to define “true positives” and “true negatives” when speaking of survival), it was not possible to compute any of the aforementioned metrics for the OS prediction. Therefore, we have developed a correctness score for the OS prediction defined for each patient, based on the true OS and the physician’s predicted OS ranges as follows:

- 0 points were assigned in case of different and non-consecutive ranges for the true OS and the predicted OS by the physician;

- 1 point was assigned in case of different, but consecutive, ranges for the true OS and the predicted OS by the physician;

- 2 points were assigned in case of identical ranges for the true OS and the predicted OS by the physician.

The Wilcoxon signed-rank test for paired data was employed to assess any statistically significant differences between the assigned scores’ distributions. A total correctness score was computed by summing all scores according to phase and physicians’ expertise.

Finally, the confusion matrix after using XAI^34^ was derived for all physicians, for experts only and non-experts only, Fig 5c and ***Supplementary Information 2***. When a medical oncologist decides with the assistance of the XAI tool, four errors can occur: (i) False Confirmation Error (FCoE), (ii) False Conflict Error (FCE), (iii) True Conflict Error (TCE), (iv) True Confirmation Error (TCoE). Furthermore, in case of correct prediction, there are 4 possible scenarios: (a) Correct True Confirmation Case (CTCo), (b) Correct True Conflict Case (CTC), (c) Correct False Confirmation Case (CFCo), (d) Correct False Conflict Case (CFC). All are summarised and explained in **Table 2**.

**Table 2.**
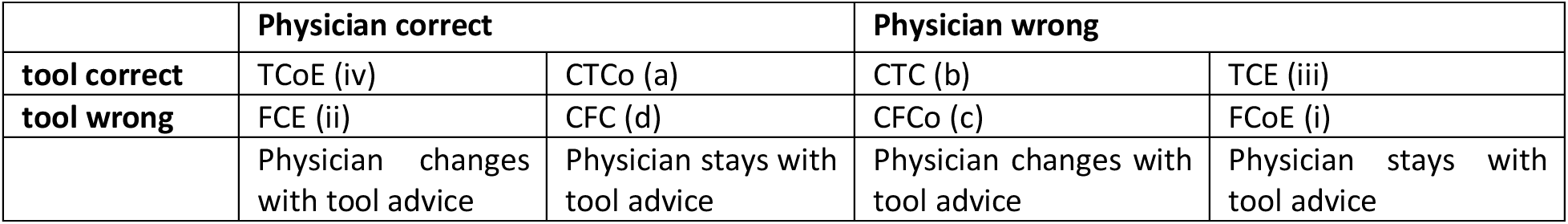
The theoretical concept behind confusion matrix of correctness of human–XAI-collaboration with regard to the physician switching or keeping the diagnosis after getting XAI advice, in which cases can fall into one of 8 scenarios: 4 errors (i–iv) and 4 correct predictions (a–d).

## Data availability

The I3LUNG project is an ongoing EU-funded initiative, with related analyses and manuscripts expected through June 2027. Therefore, the full raw multimodal dataset cannot be released at this time. However, de-identified data supporting the findings of this study, specifically, RWD and genomics data and the extracted features from medical images (radiomics, FM for CT scans and digital pathology) will be made available upon the publications on ZENODO. In line with the consortium’s data-sharing policy, the full raw datasets from all modalities (including images) will become available five years after the official end of the project.

## Code availability

The code used for preprocessing, model training, and statistical analyses is available on GitHub. For now the repository is private (https://github.com/AI-ON-Laboratory/I3LUNG_PDSS), after the acceptance, the repository will be public for everyone.

## Supplementary Information

Supplementary Information 1 - Supplementary Figures and Supplementary Tables

Supplementary Information 2 - In depth analysis of Clinical Usability Study

Supplementary Information 3 - Exploratory clinical data analysis for cohort23 by center

Supplementary Information 4 - Supporting Material for Clinical Usability Study (Global and Local SHAP tutorials and general tutorial provided to physicians on how to perform the study)

## Extended data

**Extended data Fig. 1.**
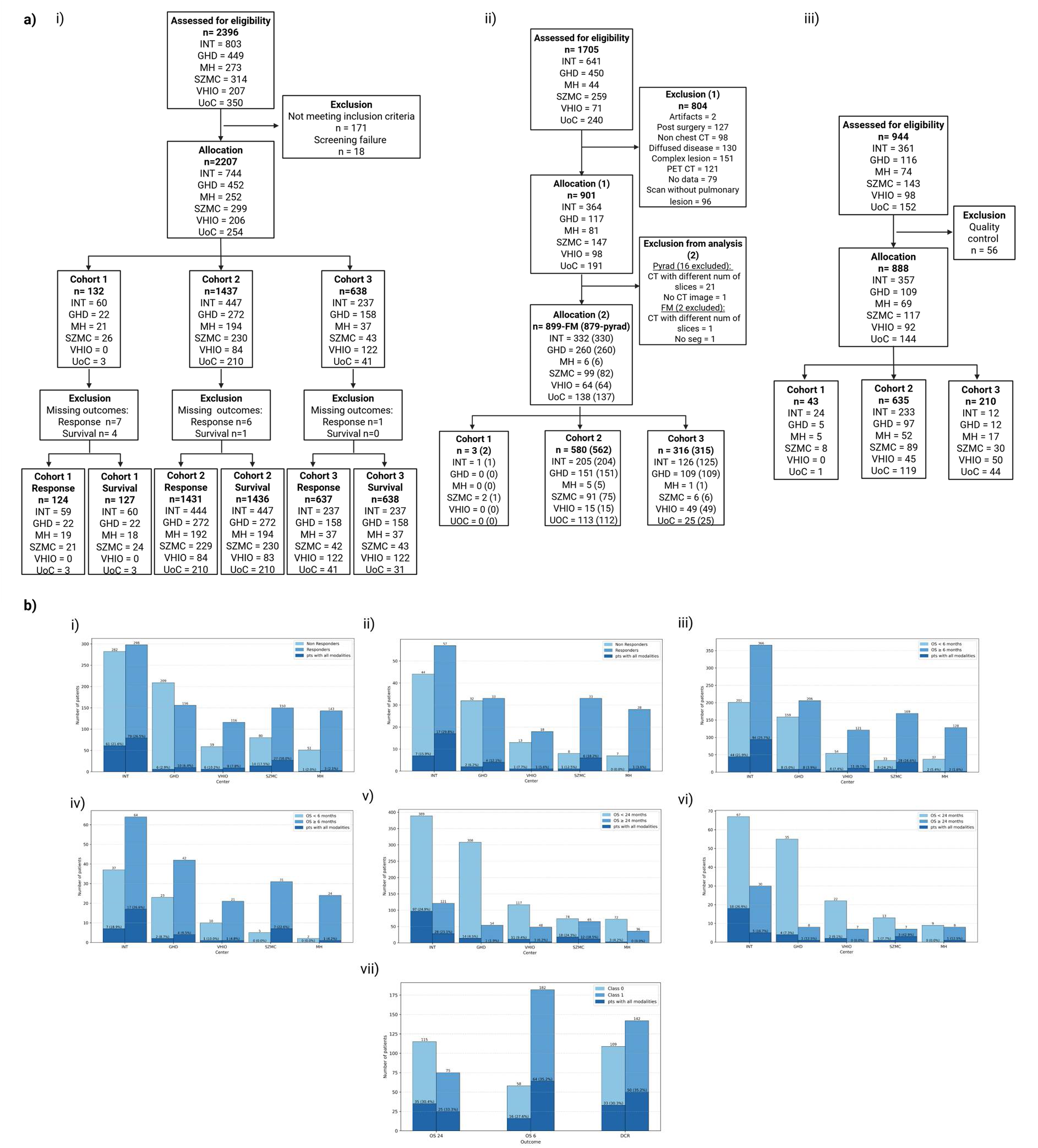
Data analysis: a) Consort flow diagram for (i) clinical data, (ii) CT images, and (iii) DP; b) Distribution of pts across centres by outcomes: DCR in TRAIN (i) and TEST (ii), OS6 in TRAIN (iii) and TEST (iv), OS24 in TRAIN (v) and TEST (vi), and EXVAL (vii)

**Extended data Fig. 2.**
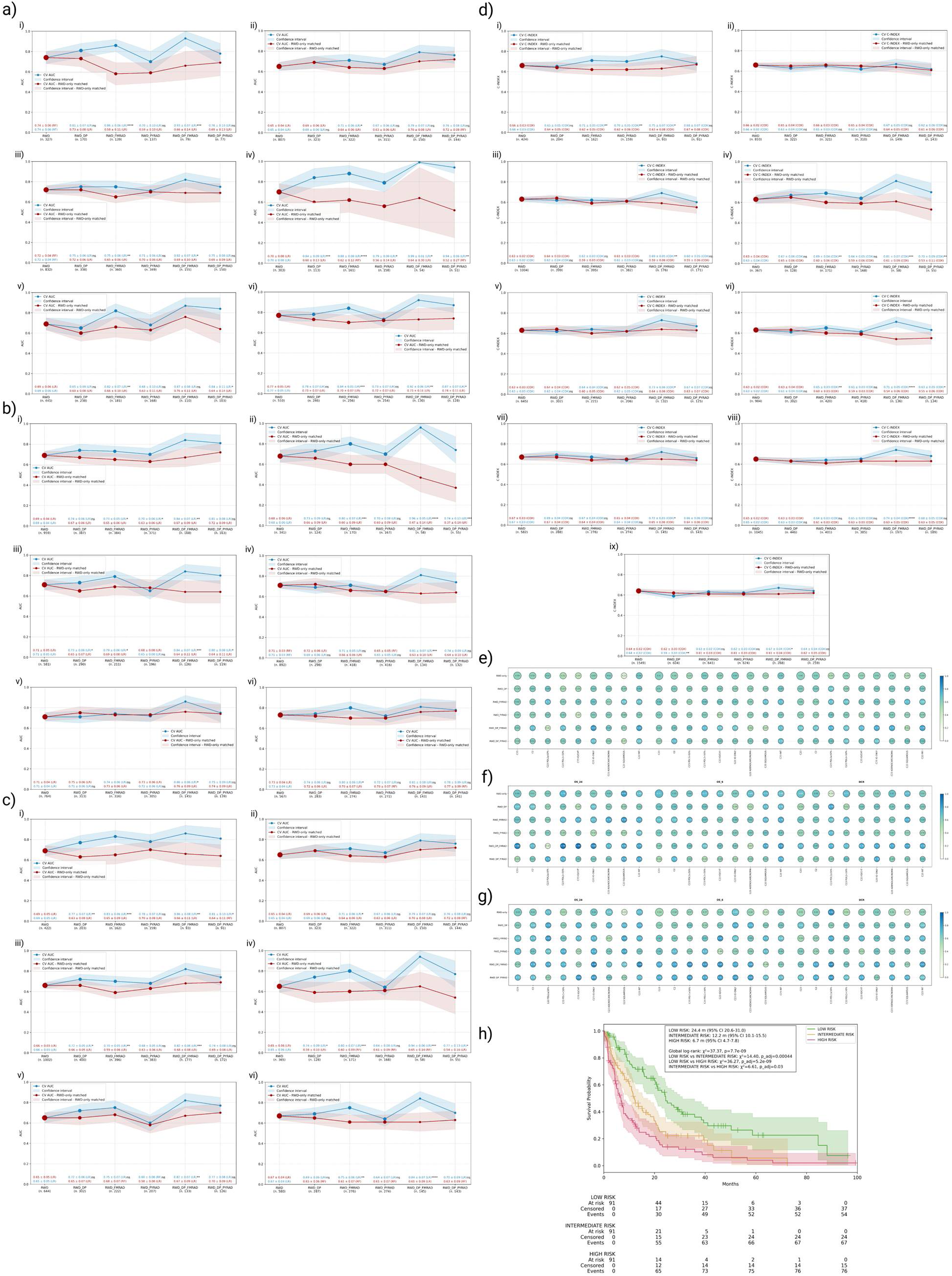
MLEF Framework: a) CV AUC for OS24 (i) C23 PDL1≥50%, (ii) C23 PDL1<50%, (iii) C23 adeno, (iv) C23 squamous, (v) C23 IO/CHT, (vi) C23 INT; b) CV AUC for OS6 (i) C23 adeno, (ii) C23 squamous, (iii) C23 IO/CHT, (iv) C23 IO-only, (v) C23 PDL1<50%, (vi) C23 INT; c) CV AUC for DCR (i) C23 PDL1≥50%, (ii) C23 PDL1<50%, (iii) C23 adeno, (iv) C23 squamous, (v) C23 IO/CHT, (vi) C23 INT; d) CV c-index for OS survival analysis results (i) C23 PDL1≥50%, (ii) C23 PDL1<50%, (iii) C23 adeno, (iv) C23 squamous, (v) C23 IO/CHT, (vi) C23 IO-only, (vii) C23 INT, (viii) C2, (ix) C23; e) CV F1 performance of MLEF classifier (OS24, OS6, DCR) across all subgroup analysis, circle color indicates the model’s performance, while size correspondence to the size of pts cohort; f) CV Sensitivity performance of MLEF classifier (OS24, OS6, DCR), circle color indicates the model’s performance, while size correspondence to the size of pts cohort; g) CV Specificity performance of MLEF classifier (OS24, OS6, DCR) across all subgroup analysis, circle color indicates the model’s performance, while size correspondence to the size of pts cohseort; h) Cox-MLEF stratification of full TEST set

**Extended data Fig. 3.**
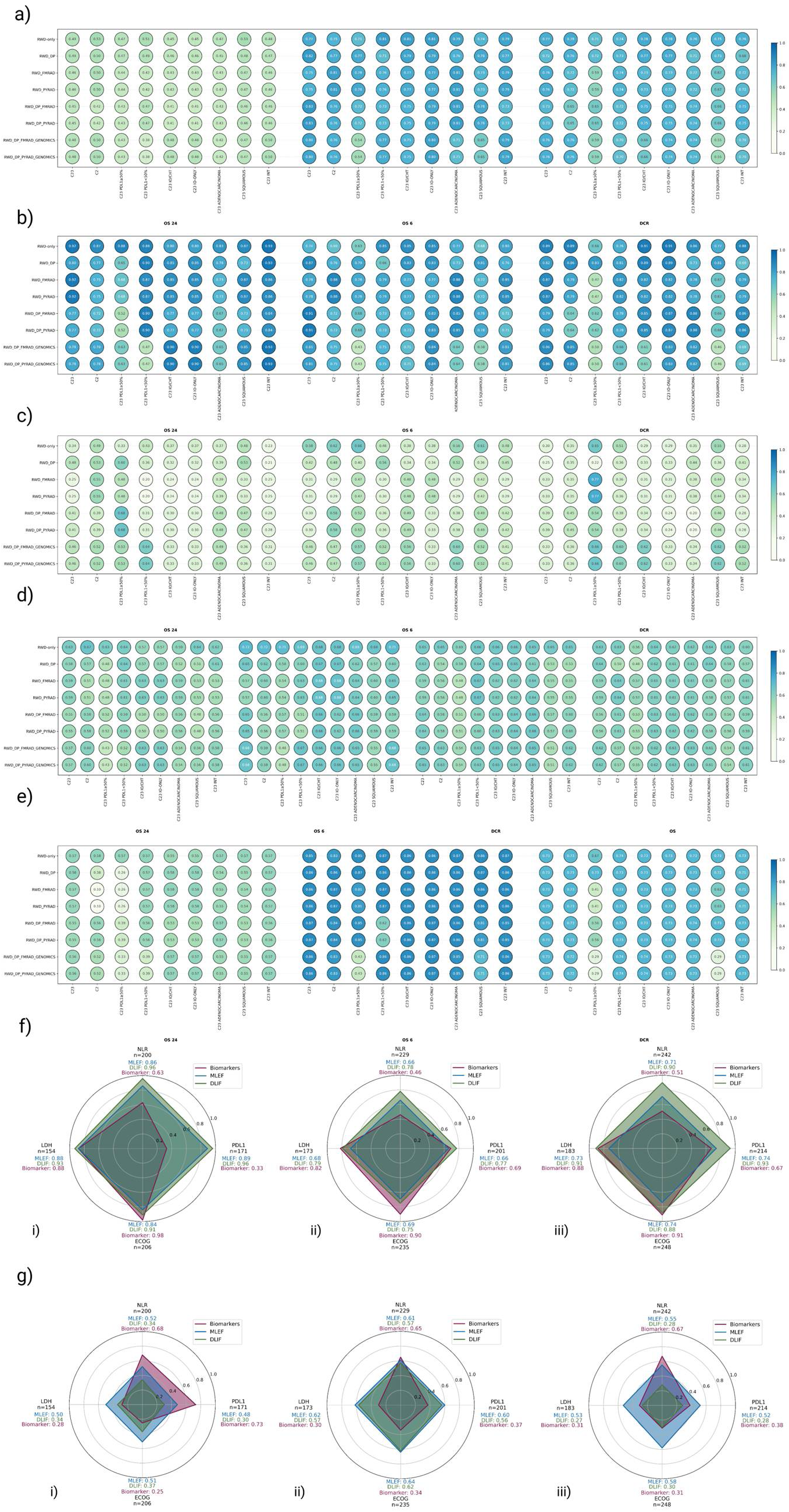
DLIF Framework: a) TEST F1 of DLIF classifier, b) TEST Sensitivity of DLIF classifier, c) TEST Specificity of DLIF classifier, d) EXVAL AUC of DLIF classifier and C-index of Cox-DLIF, e) EXVAL F1 of DLIF classifier f) Direct comparison between MLEF, DLIF and single biomarkers for (i) OS24, (ii) OS6, and (iii) DCR in the C23 cohort on TEST; performance assessed using Sensitivity g) Direct comparison between MLEF, DLIF and single biomarkers for (i) OS24, (ii) OS6, and (iii) DCR in the C23 cohort on TEST; performance assessed using Specificity, tested with DeLong’s method, p-values were calculated using a two-sided t-test.

**Extended data Fig. 4.**
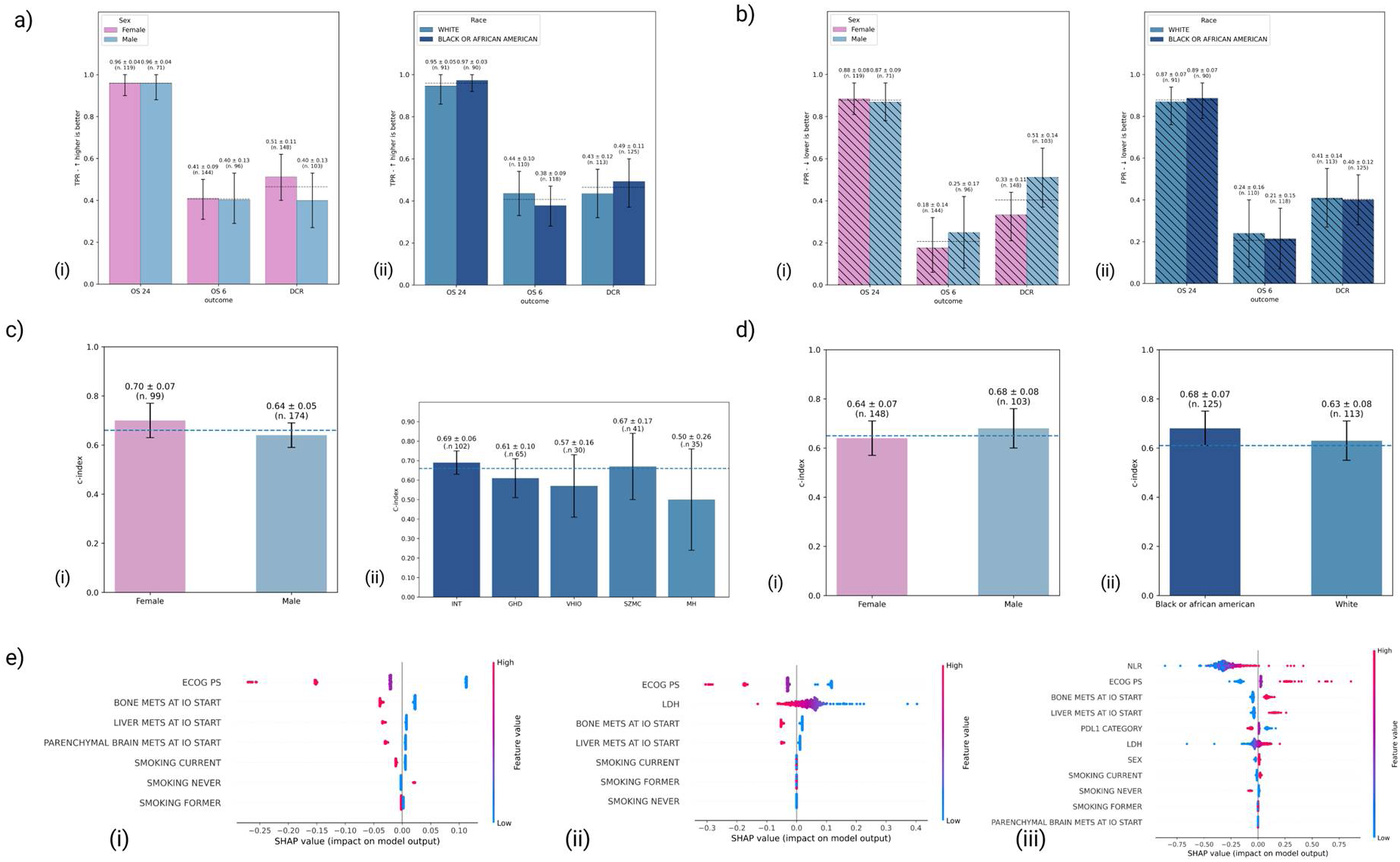
a) Fairness analysis: EXVAL TPR across sex (i) and self-reported race (ii), b) EXVAL FPR across sex (i) and self-reported race (ii), c) TEST c-index across sex (i) and centres (ii), d) EXVAL c-index across (i) self-reported race and (ii) sex ; e) post-hoc XAI analysis: global SHAP plots for OS6 (i), DCR (ii), and OS (iii)

