## Supplementary_Info_1 for "I3LUNG: Clinical Validation of a Multimodal AI Tool to Support Immunotherapy Decisions in NSCLC"

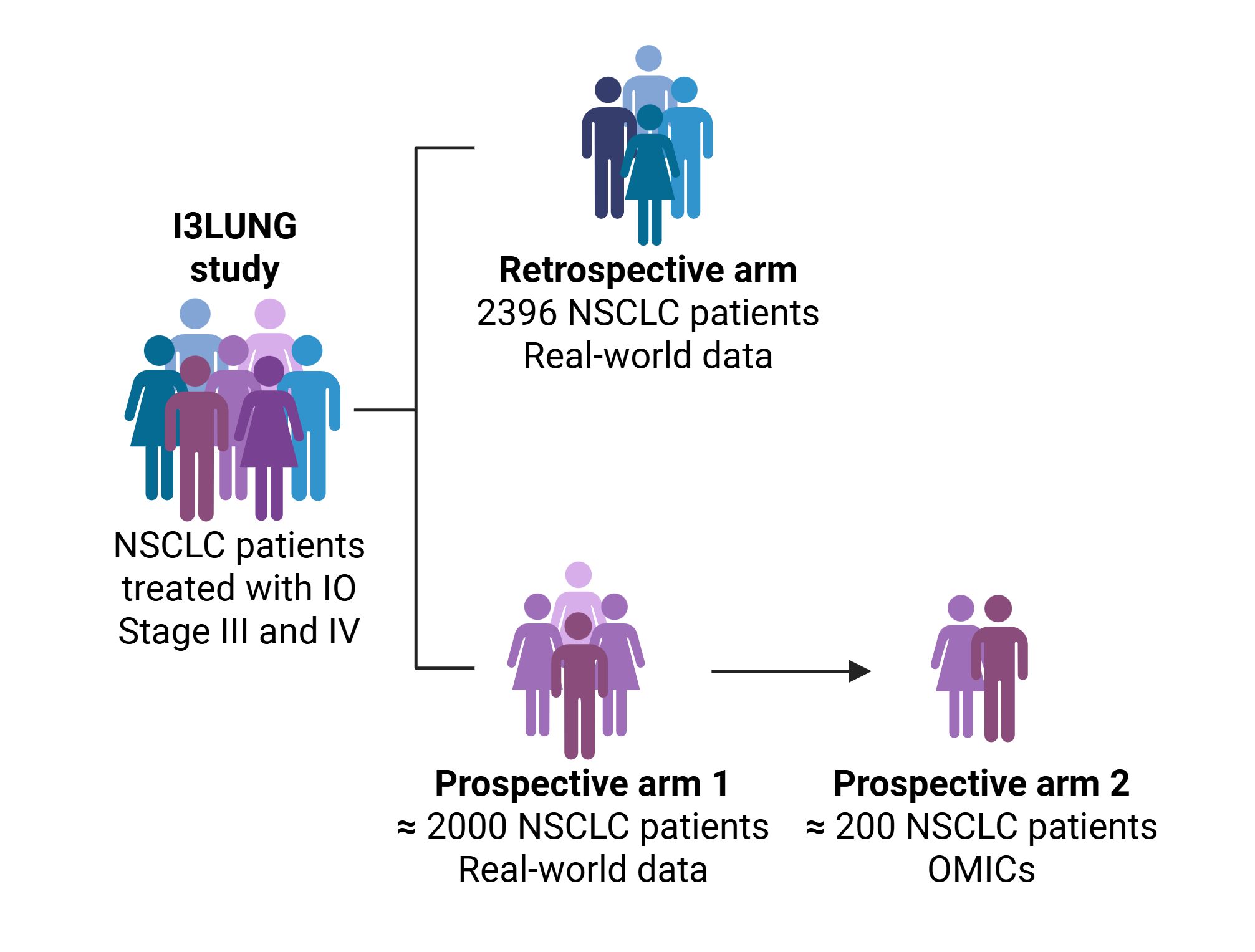

**Supplementary Fig. 1 I3LUNG study cohort overview**

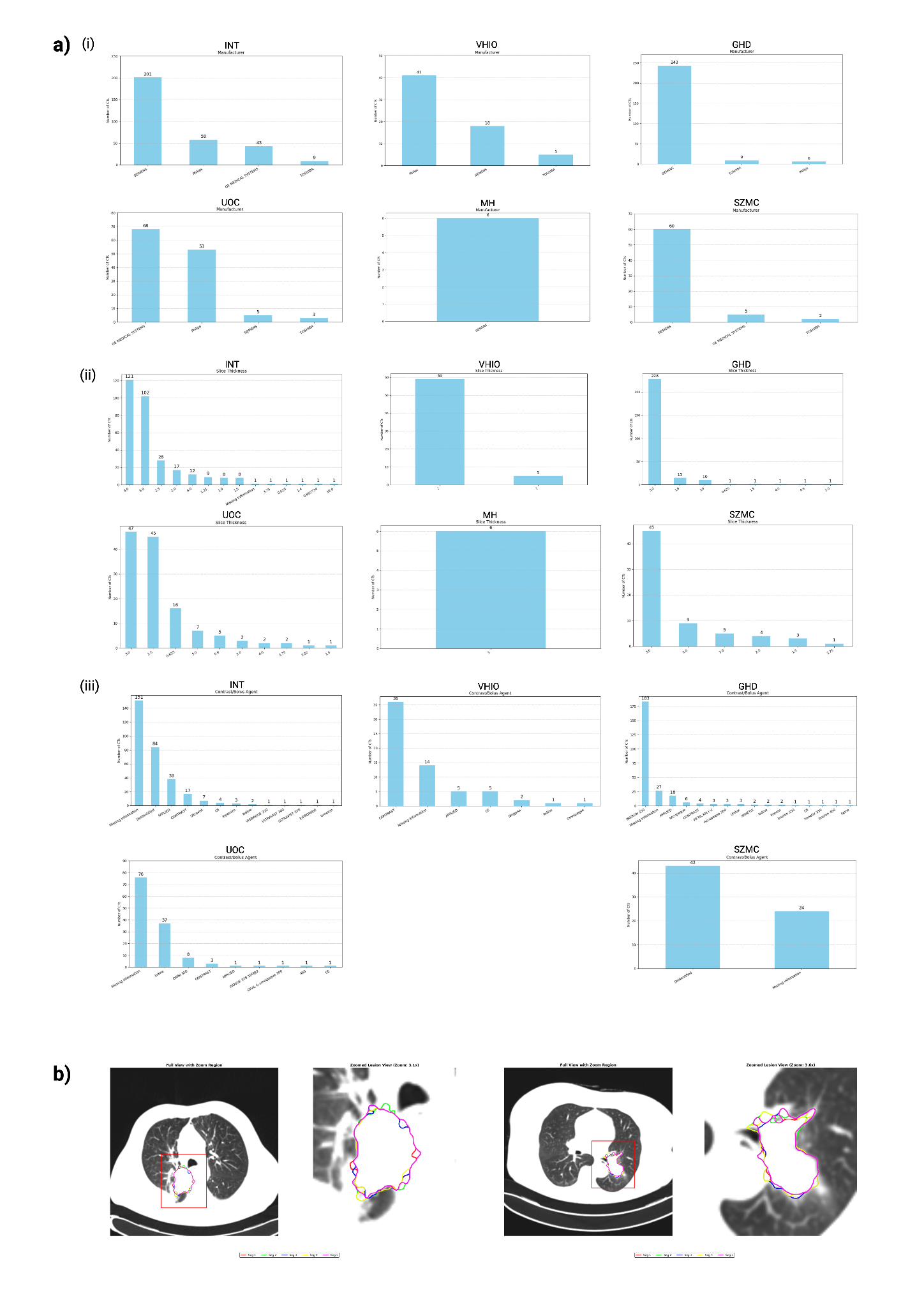

**Supplementary Fig. 2 CT scans: a) Metadata analysis (i) manufacturer (ii) slice-thickness (iii) contrast information by centre, b) example of CT segmentation with perturbations**

**Supplementary Table 1: List of subanalyses performed**

| **Name** | **Description** |
| --- | --- |
| C23 main analysis | NSCLC pts treated with IO or IO/CHT at I or further metastatic lines (Cohort 2 and 3) |
| C2 first line | NSCLC pts treated with IO or IO/CHT at I metastatic lines (Cohort 2) |
| C23 PDL1≥50% | NSCLC pts treated with IO or IO/CHT at I or further metastatic lines (Cohort 2 and 3) with PDL1≥50% |
| C23 PDL1<50% | NSCLC pts treated with IO or IO/CHT at I or further metastatic lines (Cohort 2 and 3) with PDL1<50% |
| C23 IO/CT | NSCLC pts treated with IO/CHT at I or further metastatic lines (Cohort 2 and 3) |
| C23 IO-only | NSCLC pts treated with IO-only at I or further metastatic lines (Cohort 2 and 3) |
| C23 adeno | Adenocarcinoma NSCLC pts treated with IO or IO/CHT at I or further metastatic lines (Cohort 2 and 3) |
| C23 squamous | Squamous NSCLC pts treated with IO or IO/CHT at I or further metastatic lines (Cohort 2 and 3) |
| C23 INT | NSCLC pts treated with IO or IO/CT at I or further metastatic lines (Cohort 2 and 3) from INT, Italy |

**Supplementary Table 2. Hypothesis-driven selected features and encoding**

| **Name** | **Description** | **Encoding** |
| --- | --- | --- |
| SEX | Sex of the patient | 0: female  1: male |
| ECOG | Ecog performance status | An integer between 0 and 5  0: best  5: worst |
| LDH | Lactate dehydrogenase | Numeric (U/L) |
| NLR | Neutrophils to lymphocytes ratio | Numeric |
| SMOKING_CURRENT | Current smoker | 0: no  1: yes |
| SMOKING_FORMER | Former smoker | 0: no  1: yes |
| SMOKING_NEVER | Never smoked | 0: no  1: yes |
| PDL1 | Category of PDL1 | 0: <1%  1: ≥1%, <50%  2: ≥50% |
| SITE_METS_IO_START_PARENCHYMAL BRAIN | Parenchymal brain metastases | 0: no  1: yes |
| SITE_METS_IO_START_LIVER | Liver metastases | 0: no  1: yes |
| SITE_METS_IO_START_BONE | Bone metastases | 0: no  1: yes |
| RACE* | The self-reported race of the patient | White, Asian, Native Hawaiian or other pacific islander, Black or African American, American Indian or Alaska Native |

* feature used just for fairness analysis

**Supplementary Table 3: C23 RWD-only MLEF model with full data-driven feature selection approach (input to the MLEF pipeline 227 features) MLEF-classification (a) and Cox-MLEF (b)**

a) **MLEF classification**

| **OUTCOME** | **DATASET** | **F1-macro** | **AUC** | **SENS** | **SPEC** |
| --- | --- | --- | --- | --- | --- |
| **OS24** | **CV** | 0.58 ± 0.06 | 0.68 ± 0.03 | 0.62 ± 0.05 | 0.64 ± 0.07 |
|  | **Test** | 0.62 | 0.70 ± 0.09 | 0.65 | 0.66 |
|  | **Ex. Val.** | 0.55 | 0.63 ± 0.08 | 0.45 | 0.69 |
| **OS6** | **CV** | 0.63 ± 0.05 | 0.72 ± 0.03 | 0.67 ± 0.08 | 0.61 ± 0.13 |
|  | **Test** | 0.64 | 0.70 ± 0.07 | 0.74 | 0.56 |
|  | **Ex. Val.** | 0.51 | 0.68 ± 0.08 | 0.92 | 0.12 |
| **DCR** | **CV** | 0.63 ± 0.04 | 0.71 ± 0.03 | 0.69 ± 0.13 | 0.58 ± 0.17 |
|  | **Test** | 0.66 | 0.71 ± 0.03 | 0.68 | 0.63 |
|  | **Ex. Val.** | 0.40 | 0.55 ± 0.07 | 0.18 | 0.76 |

b) **Cox-MLEF**

| **OUTCOME** | **DATASET** | **C-INDEX** |
| --- | --- | --- |
| **OS** | **CV** | 0.66 ± 0.02 |
|  | **Test** | 0.66 ± 0.04 |
|  | **Ex. Val.** | 0.65 ± 0.05 |
