## Supplementary_Info_2 for "I3LUNG: Clinical Validation of a Multimodal AI Tool to Support Immunotherapy Decisions in NSCLC"

TABLE OF CONTENTS

1. STATISTICAL METHODS	3

Table 1. Confusion matrix template.	4

2. ANALYSIS 1: PHASE 1 vs PHASE 2	5

2.1 DCR	5

Table 2. Contingency table for DCR predictions in phase 1 for all physicians.	5

Table 3. Contingency table for DCR predictions in phase 2 for all physicians.	5

Table 4. Association between DCR prediction success and XAI use for all physicians. Univariable logistic regression model.	5

Table 5. Metrics summary for DCR predictions according to phase for all physicians.	6

Table 6. Contingency table for DCR predictions in phase 1 for expert physicians.	7

Table 7. Contingency table for DCR predictions in phase 2 for expert physicians.	7

Table 8. Association between DCR prediction success and XAI use for expert physicians. Univariable logistic regression model.	7

Table 9. Metrics summary for DCR predictions according to phase for expert physicians.	8

Table 10. Contingency table for DCR predictions in phase 1 for non-expert physicians.	9

Table 11. Contingency table for DCR predictions in phase 2 for non-expert physicians.	9

Table 12. Association between DCR prediction success and XAI use for non-expert physicians. Univariable logistic regression model.	9

Table 13. Metrics summary for DCR predictions according to phase for non-expert physicians.	10

2.2 OS	11

Table 14. Contingency table for OS predictions in phase 1 for all physicians.	11

Table 15. Contingency table for OS predictions in phase 2 for all physicians.	11

Table 16. Association between OS prediction success and XAI use for all physicians. Univariable logistic regression model.	11

Table 17. Total score for OS predictions correctness for all physicians.	11

Table 18. Contingency table for OS predictions in phase 1 for expert physicians.	12

Table 19. Contingency table for OS predictions in phase 2 for expert physicians.	12

Table 20. Association between OS prediction success and XAI use for expert physicians. Univariable logistic regression model.	12

Table 21. Total score for OS predictions correctness for expert physicians.	12

Table 22. Contingency table for OS predictions in phase 1 for non-expert physicians.	13

Table 23. Contingency table for OS predictions in phase 2 for non-expert physicians.	13

Table 24. Association between OS prediction success and XAI use for non-expert physicians. Univariable logistic regression model.	13

Table 25. Total score for OS predictions correctness for non-expert physicians.	13

3. ANALYSIS 2: EXPERTS vs NON-EXPERTS (PHASE 1)	14

3.1 DCR	14

Table 26. Contingency table for DCR predictions by experts and non-experts in phase 1.	14

Table 27. Association between DCR prediction success and physicians’expertise in phase 1. Univariable logistic regression model.	14

Table 28. Metrics summary for DCR predictions according to physicians’expertise in phase 1.	14

3.2 OS	15

Table 29. Contingency table for OS predictions by experts and non-experts in phase 1.	15

Table 30. Association between OS prediction success and physicians’expertise in phase 1. Univariable logistic regression model.	15

Table 31. Total score for OS predictions according to physicians’expertise in phase 1.	15

4. ANALYSIS 3: EXPERTS vs NON-EXPERTS (PHASE 2)	16

4.1 DCR	16

Table 32. Contingency table for DCR predictions by experts and non-experts in phase 2.	16

Table 33. Association between DCR prediction success and physicians’expertise in phase 2. Univariable logistic regression model.	16

Table 34. Metrics summary for DCR predictions according to physicians’expertise in phase 2.	16

4.2 OS	17

Table 35. Contingency table for OS predictions by experts and non-experts in phase 2.	17

Table 36. Association between OS prediction success and physicians’expertise in phase 2. Univariable logistic regression model.	17

Table 37. Total score for OS predictions according to physicians’expertise in phase 2.	17

5. ANALYSIS 4: CONFUSION MATRIX	18

Table 38. Confusion matrix for DCR predictions after using the XAI for all physicians.	18

Table 39. Confusion matrix for DCR predictions after using the XAI for expert physicians.	18

Table 40. Confusion matrix for DCR predictions after using the XAI for non-expert physicians.	19

### 1. STATISTICAL METHODS

100 patients with a 1 expert to 1 non-lung expert correspondence were included in the analyses.

The association between the DCR prediction success and XAI use for all physicians, for experts only and non-lung experts only was assessed by using a univariable logistic regression model. A binary variable indicating the success of the DCR prediction (1 if the predicted DCR was equal to the true DCR, 0 otherwise) was considered as the response variable; a binary variable corresponding to the phase (1 if phase 1/no XAI use, 2 if phase 2/XAI use) was treated as the covariate. The results were presented as Odds Ratio (OR) and Area Under the Curve (AUC) with their 95% Confidence Interval (95% CI).

The associations between the DCR prediction success and physicians‘expertise in phase 1 and phase 2 were assessed by using a univariable logistic regression model. A binary variable indicating the success of the DCR prediction (1 if the predicted DCR was equal to the true DCR, 0 otherwise) was considered as the response variable; a binary variable corresponding to the expertise level (0 if non-expert, 1 if expert) was treated as the covariate. The results were presented as Odds Ratio (OR) and Area Under the Curve (AUC) with their 95% Confidence Interval (95% CI).

The association between the OS prediction success and XAI use for all physicians, for experts only and non-experts only was assessed by using a univariable logistic regression model. A binary variable indicating the success of the OS prediction (1 if the [true OS - 25%; true OS + 25%] interval overlapped with the predicted OS range, 0 otherwise) was considered as the response variable; a binary variable corresponding to the phase (1 if phase 1/no XAI use, 2 if phase 2/XAI use) was treated as the covariate. The results were presented as Odds Ratio (OR) and Area Under the Curve (AUC) with their 95% Confidence Interval (95% CI).

The associations between the OS prediction success and physicians‘expertise in phase 1 and phase 2 were assessed by using a univariable logistic regression model. A binary variable indicating the success of the OS prediction (1 if the [true OS - 25%; true OS + 25%] interval overlapped with the predicted OS range, 0 otherwise) was considered as the response variable; a binary variable corresponding to the expertise level (0 if non-expert, 1 if expert) was treated as the covariate. The results were presented as Odds Ratio (OR) and Area Under the Curve (AUC) with their 95% Confidence Interval (95% CI).

The degree of agreement between the expert and non-expert physicians’ DCR and OS evaluations were assessed by computing the Cohen’s Kappa and the weighted Cohen’s Kappa, respectively. Based on the observed Kappa, the degree of concordance were considered as follows:

- 0: no agreement

- [0.01 - 0.20]: slight agreement,

- [0.21 - 0.40]: fair agreement,

- [0.41 - 0.60]: moderate agreement,

- [0.61 - 0.80]: substantial agreement,

- [0.81 - 0.99]: near perfect agreement,

- 1: perfect agreement.

The McNemar’s test for paired data was employed to assess any differences between the experts and the non-experts in the discordant predictions in phase 1 and phase 2.

Furthermore, metrics such as the sensitivity, specificity, accuracy and precision were computed for the DCR prediction according to phase and physicians’expertise. The 95% CIs of these metrics were calculated by using the Clopper-Pearson’s exact binomial method. The McNemar’s test for paired data was employed to assess any statistically significant differences.

The F1-score was derived according to phase and physicians’expertise for the DCR prediction. Its 95% CI was computed by using bootstrap methods with 1000 replications.

Due to the time-to-event nature of the OS endpoint (i.e., it is not possible to define “true positives” and “true negatives” when speaking of survival) it is not possible to compute any of the aforementioned metrics for the OS prediction.

A correctness score for the OS prediction was defined for each patient based on the true OS and the physician’s predicted OS ranges as follows:

- A score of 0 points was assigned in case of different and non-consecutive ranges for the true OS and the predicted OS by the physician;

- A score of 1 point was assigned in case of different, but consecutive, ranges for the true OS and the predicted OS by the physician;

- A score of 2 points was assigned in case of identical ranges for the true OS and the predicted OS by the physician.

The Wilcoxon signed-rank test for paired data was employed to assess any statistically significant differences between the assigned scores’distributions. A total correctness score was computed by summing all scores according to the phase and physicians’expertise.

Finally, the confusion matrix after using the XAI was derived according to Rosenbacke et al. for all physicians, for experts only and non-experts only.

The following table shows the confusion matrix template:

##### Table 1. Confusion matrix template.

|  | **Physician correct** |  | **Physician wrong** |  |
| --- | --- | --- | --- | --- |
| **XAI correct** | True confirmation error | True confirmation (correct) | True conflict (correct) | True conflict error |
| **XAI wrong** | False conflict error | False conflict (correct) | False confirmation (correct) | False confirmation error |
|  | *Physician changes after XAI advice* | *Physician stays after XAI advice* | *Physician changes after XAI advice* | *Physician stays after XAI advice* |

### 2. ANALYSIS 1: PHASE 1 vs PHASE 2

#### 2.1 DCR

##### Table 2. Contingency table for DCR predictions in phase 1 for all physicians.

| **Predicted DCR** | **DCR (Ground truth)** | |
| --- | --- | --- |
|  | **PD** | **DCR** |
| **PD** | 27 (13.5%) | 33 (16.5%) |
| **DCR** | 53 (26.5%) | 87 (43.5%) |

In the 57% of cases the predicted DCR coincides with the ground truth (13.5%+43.5%) before using the XAI (all physicians).

##### Table 3. Contingency table for DCR predictions in phase 2 for all physicians.

| **Predicted DCR** | **DCR (Ground truth)** | |
| --- | --- | --- |
|  | **PD** | **DCR** |
| **PD** | 25 (12.5%) | 16 (8.0%) |
| **DCR** | 55 (27.5%) | 104 (52.0%) |

In the 62.5% of cases the predicted DCR coincides with the ground truth (12.5%+52.0%) after using the XAI (all physicians).

##### Table 4. Association between DCR prediction success and XAI use for all physicians. Univariable logistic regression model.

| **N = 400** |  |  |
| --- | --- | --- |
| **Phase** |  | p = 0.1250 |
| 1 | reference |  |
| 2 | OR (95% CI) = 1.37 (0.92-2.05) | p = 0.1250 |
| **AUC (95% CI)** | 0.539 (0.489-0.590) |  |
| **Legend:** N: number of predictions. OR: Odds Ratio. AUC: Area Under the Curve. CI: Confidence Interval. | | |

**NOTE:** ORR = 1.37 means that, overall, using the XAI increases the probability of success (i.e., predicting the true DCR) by 37%. However, this association is not statistically significant.

##### Table 5. Metrics summary for DCR predictions according to phase for all physicians.

|  | **Phase 1** | **Phase 2** | **p-value** |
| --- | --- | --- | --- |
| **Sensitivity (95% CI)** | 0.725 (0.636-0.803) | 0.867 (0.793-0.922) | 0.0011 |
| **Specificity (95% CI)** | 0.338 (0.236-0.452) | 0.313 (0.214-0.426) | 0.7055 |
| **Accuracy (95% CI)** | 0.570 (0.498-0.640) | 0.645 (0.574-0.711) | 0.0431 |
| **Precision (95% CI)** | 0.621 (0.536-0.702) | 0.654 (0.575-0.728) | 0.2498 |
| **F1-Score (95% CI)** | 0.669 (0.598-0.733) | 0.746 (0.687-0.803) | - |

We can reasonably infer that the sensitivity improved after the XAI use based on the non-inclusion in each other’s 95% CI. This means that using the XAI changes the physician's opinion on false negatives and it increases the number of DCR patients to be treated. Based on the same interpretation, using the XAI also improves the accuracy (the overall percentage of correct evaluations) and the F1-score. The McNemar’s test confirms the conclusions made on the sensitivity and accuracy, since the differences are statistically significant at the 5% nominal level.

##### Table 6. Contingency table for DCR predictions in phase 1 for expert physicians.

| **Predicted DCR** | **DCR (Ground truth)** | |
| --- | --- | --- |
|  | **PD** | **DCR** |
| **PD** | 17 (17.0%) | 19 (19.0%) |
| **DCR** | 23 (23.0%) | 41 (41.0%) |

In the 58% of cases the predicted DCR coincides with the ground truth (17.0%+41.0%) before using XAI (experts only).

##### Table 7. Contingency table for DCR predictions in phase 2 for expert physicians.

| **Predicted DCR** | **DCR (Ground truth)** | |
| --- | --- | --- |
|  | **PD** | **DCR** |
| **PD** | 13 (13.0%) | 10 (10.0%) |
| **DCR** | 27 (27.0%) | 50 (50.0%) |

In the 63% of cases the predicted DCR coincides with the ground truth (13.0%+50.0%) after using XAI (experts only).

##### Table 8. Association between DCR prediction success and XAI use for expert physicians. Univariable logistic regression model.

| **N = 200** |  |  |
| --- | --- | --- |
| **Phase** |  | p = 0.4698 |
| 1 | reference |  |
| 2 | OR (95% CI) = 1.23 (0.70-2.18) | p = 0.4698 |
| **AUC (95% CI)** | 0.526 (0.455-0.597) |  |
| **Legend:** N: number of predictions. OR: Odds Ratio. AUC: Area Under the Curve. CI: Confidence Interval. | | |

**NOTE:** ORR = 1.23 means that, for expert physicians, using the XAI increases the probability of success (i.e., predicting the true DCR) by 23%. However, this association is not statistically significant.

##### Table 9. Metrics summary for DCR predictions according to phase for expert physicians.

|  | **Phase 1** | **Phase 2** | **p-value** |
| --- | --- | --- | --- |
| **Sensitivity (95% CI)** | 0.683 (0.550-0.797) | 0.833 (0.715-0.917) | 0.0201 |
| **Specificity (95% CI)** | 0.425 (0.270-0.591) | 0.325 (0.186-0.491) | 0.3173 |
| **Accuracy (95% CI)** | 0.580 (0.477-0.678) | 0.630 (0.528-0.724) | 0.3692 |
| **Precision (95% CI)** | 0.641 (0.511-0.757) | 0.649 (0.532-0.755) | 0.6698 |
| **F1-Score (95% CI)** | 0.661 (0.560-0.752) | 0.730 (0.641-0.808) | - |

We can reasonably infer that the sensitivity improved after the XAI use based on the non-inclusion in each other’s 95% CI. This means that using the XAI changes the expert's opinion on false negatives and it increases the number of DCR patients to be treated. The McNemar’s test confirms the conclusions made on the sensitivity, since the difference is statistically significant at the 5% nominal level.

##### Table 10. Contingency table for DCR predictions in phase 1 for non-expert physicians.

| **Predicted DCR** | **DCR (Ground truth)** | |
| --- | --- | --- |
|  | **PD** | **DCR** |
| **PD** | 10 (10.0%) | 14 (14.0%) |
| **DCR** | 30 (30.0%) | 46 (46.0%) |

In the 56% of cases the predicted DCR coincides with the ground truth before using the XAI (non-experts only).

##### Table 11. Contingency table for DCR predictions in phase 2 for non-expert physicians.

| **Predicted DCR** | **DCR (Ground truth)** | |
| --- | --- | --- |
|  | **PD** | **DCR** |
| **PD** | 12 (12.0%) | 6 (6.0%) |
| **DCR** | 28 (28.0%) | 54 (54.0%) |

In the 66% of cases the predicted DCR coincides with the ground truth after using the XAI (non-experts only).

##### Table 12. Association between DCR prediction success and XAI use for non-expert physicians. Univariable logistic regression model.

| **N = 200** |  |  |
| --- | --- | --- |
| **Phase** |  | p = 0.1480 |
| 1 | reference |  |
| 2 | OR (95% CI) = 1.53 (0.86-2.60) | p = 0.1480 |
| **AUC (95% CI)** | 0.553 (0.482-0.624) |  |
| **Legend:** N: number of predictions. OR: Odds Ratio. AUC: Area Under the Curve. CI: Confidence Interval. | | |

**NOTE:** ORR = 1.53 means that, for non-expert physicians, using the XAI increases the probability of success (i.e., predicting the true DCR) by 53%. However, this association is not statistically significant.

##### Table 13. Metrics summary for DCR predictions according to phase for non-expert physicians.

|  | **Phase 1** | **Phase 2** | **p-value** |
| --- | --- | --- | --- |
| **Sensitivity (95% CI)** | 0.767 (0.640-0.866) | 0.900 (0.795-0.962) | 0.0209 |
| **Specificity (95% CI)** | 0.250 (0.127-0.412) | 0.300 (0.166-0.465) | 0.5637 |
| **Accuracy (95% CI)** | 0.560 (0.457-0.659) | 0.660 (0.559-0.752) | 0.0412 |
| **Precision (95% CI)** | 0.605 (0.487-0.716) | 0.659 (0.546-0.760) | 0.1967 |
| **F1-Score (95% CI)** | 0.676 (0.583-0.758) | 0.761 (0.678-0.830) | - |

We can reasonably infer that the sensitivity improved after the XAI use based on the non-inclusion in each other’s 95% CI. This means that using the XAI changes the non-expert's opinion on false negatives and it increases the number of DCR patients to be treated. Based on the same interpretation, using the XAI also improves the accuracy (the overall percentage of correct evaluations) and the F1-score. The McNemar’s test confirms the conclusions made on the sensitivity and the accuracy, since the differences are statistically significant.

## 2.2 OS

##### Table 14. Contingency table for OS predictions in phase 1 for all physicians.

| **Predicted OS** | **OS (Ground truth)** | | | | |
| --- | --- | --- | --- | --- | --- |
|  | **< 6 months** | **[6-12) months** | **[12-18) months** | **[18-24) months** | **≥24 months** |
| **< 6 months** | 4 (2.0%) | 5 (2.5%) | 2 (1.0%) | 0 (0.0%) | 4 (2.0%) |
| **[6-12) months** | 20 (10.0%) | 12 (6.0%) | 8 (4.0%) | 9 (4.5%) | 18 (9.0%) |
| **[12-18) months** | 13 (6.5%) | 10 (5.0%) | 7 (3.5%) | 6 (3.0%) | 21 (10.5%) |
| **[18-24) months** | 9 (4.5%) | 10 (5.0%) | 3 (1.5%) | 1 (0.5%) | 14 (7.0%) |
| **≥24 months** | 10 (5.0%) | 1 (0.5%) | 2 (1.0%) | 2 (1.0%) | 9 (4.5%) |

##### Table 15. Contingency table for OS predictions in phase 2 for all physicians.

| **Predicted OS** | **OS (Ground truth)** | | | | |
| --- | --- | --- | --- | --- | --- |
|  | **< 6 months** | **[6-12) months** | **[12-18) months** | **[18-24) months** | **≥24 months** |
| **< 6 months** | 8 (4.0%) | 1 (0.5%) | 1 (0.5%) | 3 (1.5%) | 1 (0.5%) |
| **[6-12) months** | 16 (8.0%) | 15 (7.5%) | 8 (4.0%) | 3 (1.5%) | 13 (6.5%) |
| **[12-18) months** | 17 (8.5%) | 11 (5.5%) | 4 (2.0%) | 9 (4.5%) | 24 (12.0%) |
| **[18-24) months** | 6 (3.0%) | 8 (4.0%) | 4 (2.0%) | 2 (1.0%) | 12 (6.0%) |
| **≥24 months** | 9 (4.5%) | 3 (1.5%) | 5 (2.5%) | 1 (0.5%) | 16 (8.0%) |

##### Table 16. Association between OS prediction success and XAI use for all physicians. Univariable logistic regression model.

| **N = 400** |  |  |
| --- | --- | --- |
| **Phase** |  | p = 0.1440 |
| 1 | reference |  |
| 2 | OR (95% CI) = 1.36 (0.90-2.05) | p = 0.1440 |
| **AUC (95% CI)** | 0.538 (0.487-0.589) |  |
| **Legend:** N: number of predictions. OR: Odds Ratio. AUC: Area Under the Curve. CI: Confidence Interval. | | |

**NOTE:** ORR = 1.36 means that, overall, using the XAI increases the probability of success (i.e., predicting the true OS) by 36%. However, this association is not statistically significant.

##### Table 17. Total score for OS predictions correctness for all physicians.

| **Phase 1** | **Phase 2** | **Wilcoxon signed-rank test, p-value** |
| --- | --- | --- |
| 134/400 (33.5%) | 152/400 (38.0%) | 0.1149 |

Using the XAI appears to improve the correctness of the OS prediction. However, from the Wilcoxon signed-rank test, no statistical differences between the scores’distributions were found at the 5% level, hence no definitive conclusion can be made about this improvement of the total score between phase 1 and phase 2.

##### Table 18. Contingency table for OS predictions in phase 1 for expert physicians.

| **Predicted OS** | **OS (Ground truth)** | | | | |
| --- | --- | --- | --- | --- | --- |
|  | **< 6 months** | **[6-12) months** | **[12-18) months** | **[18-24) months** | **≥24 months** |
| **< 6 months** | 1 (1.0%) | 2 (2.0%) | 1 (1.0%) | 0 (0.0%) | 1 (0.0%) |
| **[6-12) months** | 13 (13.0%) | 6 (6.0%) | 5 (5.0%) | 5 (5.0%) | 7 (7.0%) |
| **[12-18) months** | 5 (5.0%) | 6 (6.0%) | 3 (3.0%) | 2 (2.0%) | 11 (11.0%) |
| **[18-24) months** | 4 (4.0%) | 5 (5.0%) | 2 (2.0%) | 0 (0.0%) | 8 (8.0%) |
| **≥24 months** | 5 (5.0%) | 5 (0.0%) | 2 (2.0%) | 2 (2.0%) | 6 (6.0%) |

##### Table 19. Contingency table for OS predictions in phase 2 for expert physicians.

| **Predicted OS** | **OS (Ground truth)** | | | | |
| --- | --- | --- | --- | --- | --- |
|  | **< 6 months** | **[6-12) months** | **[12-18) months** | **[18-24) months** | **≥24 months** |
| **< 6 months** | 4 (4.0%) | 1 (1.0%) | 0 (0.0%) | 1 (1.0%) | 1 (1.0%) |
| **[6-12) months** | 8 (8.0%) | 7 (7.0%) | 4 (4.0%) | 1 (1.0%) | 7 (7.0%) |
| **[12-18) months** | 9 (9.0%) | 5 (5.0%) | 2 (2.0%) | 5 (5.0%) | 11 (11.0%) |
| **[18-24) months** | 5 (5.0%) | 6 (6.0%) | 3 (3.0%) | 1 (1.0%) | 6 (6.0%) |
| **≥24 months** | 2 (2.0%) | 0 (0.0%) | 1 (1.0%) | 1 (1.0%) | 8 (8.0%) |

##### Table 20. Association between OS prediction success and XAI use for expert physicians. Univariable logistic regression model.

| **N = 200** |  |  |
| --- | --- | --- |
| **Phase** |  | p = 0.6555 |
| 1 | reference |  |
| 2 | OR (95% CI) = 1.14 (0.64-2.05) | p = 0.6555 |
| **AUC (95% CI)** | 0.517 (0.443-0.590) |  |
| **Legend:** N: number of predictions. OR: Odds Ratio. AUC: Area Under the Curve. CI: Confidence Interval. | | |

**NOTE:** ORR = 1.14 means that, overall, using the XAI increases the probability of success (i.e., predicting the true OS) by 14%. However, this association is not statistically significant.

##### Table 21. Total score for OS predictions correctness for expert physicians.

| **Phase 1** | **Phase 2** | **Wilcoxon signed-rank test, p-value** |
| --- | --- | --- |
| 72/200 (36.0%) | 77/200 (38.5%) | 0.5491 |

Using the XAI appears to improve the correctness of the OS prediction. However, from the Wilcoxon signed-rank test, no statistical differences between the scores’distributions were found at the 5% level, hence no definitive conclusion can be made about this improvement of the total score between phase 1 and phase 2.

##### Table 22. Contingency table for OS predictions in phase 1 for non-expert physicians.

| **Predicted OS** | **OS (Ground truth)** | | | | |
| --- | --- | --- | --- | --- | --- |
|  | **< 6 months** | **[6-12) months** | **[12-18) months** | **[18-24) months** | **≥24 months** |
| **< 6 months** | 3 (3.0%) | 3 (3.0%) | 1 (1.0%) | 0 (0.0%) | 3 (3.0%) |
| **[6-12) months** | 7 (7.0%) | 6 (6.0%) | 3 (3.0%) | 4 (4.0%) | 11 (11.0%) |
| **[12-18) months** | 8 (8.0%) | 4 (4.0%) | 4 (3.0%) | 4 (4.0%) | 10 (10.0%) |
| **[18-24) months** | 5 (5.0%) | 5 (5.0%) | 1 (1.0%) | 1 (1.0%) | 6 (6.0%) |
| **≥24 months** | 5 (5.0%) | 1 (1.0%) | 2 (2.0%) | 0 (0.0%) | 3 (3.0%) |

##### Table 23. Contingency table for OS predictions in phase 2 for non-expert physicians.

| **Predicted OS** | **OS (Ground truth)** | | | | |
| --- | --- | --- | --- | --- | --- |
|  | **< 6 months** | **[6-12) months** | **[12-18) months** | **[18-24) months** | **≥24 months** |
| **< 6 months** | 4 (4.0%) | 0 (0.0%) | 1 (1.0%) | 2 (2.0%) | 0 (0.0%) |
| **[6-12) months** | 8 (8.0%) | 8 (8.0%) | 4 (4.0%) | 2 (2.0%) | 6 (6.0%) |
| **[12-18) months** | 8 (8.0%) | 6 (6.0%) | 2 (2.0%) | 4 (4.0%) | 13 (13.0%) |
| **[18-24) months** | 1 (1.0%) | 2 (2.0%) | 1 (1.0%) | 1 (1.0%) | 6 (6.0%) |
| **≥24 months** | 7 (7.0%) | 3 (3.0%) | 3 (3.0%) | 0 (0.0%) | 8 (8.0%) |

##### Table 24. Association between OS prediction success and XAI use for non-expert physicians. Univariable logistic regression model.

| **N = 200** |  |  |
| --- | --- | --- |
| **Phase** |  | p = 0.1072 |
| 1 | reference |  |
| 2 | OR (95% CI) = 1.61 (0.90-2.88) | p = 0.1072 |
| **AUC (95% CI)** | 0.559 (0.488-0.631) |  |
| **Legend:** N: number of predictions. OR: Odds Ratio. AUC: Area Under the Curve. CI: Confidence Interval. | | |

**NOTE:** ORR = 1.61 means that, overall, using the XAI increases the probability of success (i.e., predicting the true OS) by 61%. However, this association is not statistically significant.

##### Table 25. Total score for OS predictions correctness for non-expert physicians.

| **Phase 1** | **Phase 2** | **Wilcoxon signed-rank test, p-value** |
| --- | --- | --- |
| 62/200 (31.0%) | 75/200 (37.5%) | 0.0759 |

Using the XAI appears to improve the correctness of the OS prediction. However, from the Wilcoxon signed-rank test, no statistical differences between the scores’distributions were found at the 5% level, hence no definitive conclusion can be made about this improvement of the total score between phase 1 and phase 2.

### 3. ANALYSIS 2: EXPERTS vs NON-EXPERTS (PHASE 1)

#### 3.1 DCR

##### Table 26. Contingency table for DCR predictions by experts and non-experts in phase 1.

| **Predicted DCR by experts** | **Predicted DCR by non-experts** | |
| --- | --- | --- |
|  | **PD** | **DCR** |
| **PD** | 11 (11.0%) | 25 (25.0%) |
| **DCR** | 13 (13.0%) | 51 (51.0%) |
| **Cohen’s Kappa** | 0.1105 (slight agreement) | |
| **McNemar’s test p-value** | 0.0516 (no significant difference between discordant predictions) | |

##### Table 27. Association between DCR prediction success and physicians’expertise in phase 1. Univariable logistic regression model.

| **N = 200** |  |  |
| --- | --- | --- |
| **Level of expertise** |  | p = 0.7752 |
| Non-expert | reference |  |
| Expert | OR (95% CI) = 1.09 (0.62-1.90) | p = 0.7752 |
| **AUC (95% CI)** | 0.510 (0.440-0.581) |  |
| **Legend:** N: number of predictions. OR: Odds Ratio. AUC: Area Under the Curve. CI: Confidence Interval. | | |

**NOTE:** ORR = 1.09 means that being an expert increases the probability of success (i.e., predicting the true DCR) by 9% in phase 1. However, this association is not statistically significant.

##### Table 28. Metrics summary for DCR predictions according to physicians’expertise in phase 1.

|  | **Non-expert** | **Expert** | **p-value** |
| --- | --- | --- | --- |
| **Sensitivity (95% CI)** | 0.767 (0.640-0.866) | 0.683 (0.550-0.797) | 0.2752 |
| **Specificity (95% CI)** | 0.250 (0.127-0.412) | 0.425 (0.270-0.591) | 0.0896 |
| **Accuracy (95% CI)** | 0.560 (0.457-0.659) | 0.580 (0.477-0.678) | 0.7456 |
| **Precision (95% CI)** | 0.605 (0.487-0.716) | 0.641 (0.511-0.757) | 0.4054 |
| **F1-Score (95% CI)** | 0.676 (0.583-0.758) | 0.661 (0.560-0.752) | - |

Before using the XAI, the experts appear to present a better specificity based on the non-inclusion in each other’s 95% CI. This means that an expert was better in identifying PD patients who should not be treated. However, from the McNemar test, this difference is not statistically significant.

## 3.2 OS

##### Table 29. Contingency table for OS predictions by experts and non-experts in phase 1.

| **Predicted OS by experts** | **Predicted OS by non-experts** | | | | |
| --- | --- | --- | --- | --- | --- |
|  | **< 6 months** | **[6-12) months** | **[12-18) months** | **[18-24) months** | **≥24 months** |
| **< 6 months** | 1 (1.0%) | 2 (2.0%) | 2 (2.0%) | 0 (0.0%) | 0 (0.0%) |
| **[6-12) months** | 4 (4.0%) | 10 (10.0%) | 13 (13.0%) | 7 (7.0%) | 2 (2.0%) |
| **[12-18) months** | 2 (2.0%) | 13 (13.0%) | 6 (6.0%) | 5 (5.0%) | 1 (1.0%) |
| **[18-24) months** | 1 (1.0%) | 4 (4.0%) | 6 (6.0%) | 5 (5.0%) | 3 (3.0%) |
| **≥24 months** | 2 (2.0%) | 2 (2.0%) | 3 (3.0%) | 1 (1.0%) | 5 (5.0%) |
| **Cohen’s Weighted Kappa** | 0.1426 (slight agreement) | | | | |

##### Table 30. Association between OS prediction success and physicians’expertise in phase 1. Univariable logistic regression model.

| **N = 200** |  |  |
| --- | --- | --- |
| **Level of expertise** |  | p = 0.7618 |
| Non-expert | reference |  |
| Expert | OR (95% CI) = 1.10 (0.61-1.99) | p = 0.7618 |
| **AUC (95% CI)** | 0.512 (0.437-0.586) |  |
| **Legend:** N: number of predictions. OR: Odds Ratio. AUC: Area Under the Curve. CI: Confidence Interval. | | |

**NOTE:** ORR = 1.10 means that being an expert increases the probability of success (i.e., predicting the true OS) by 10% in phase 1. However, this association is not statistically significant.

##### Table 31. Total score for OS predictions according to physicians’expertise in phase 1.

| **Non-expert** | **Expert** | **Wilcoxon signed-rank test, p-value** |
| --- | --- | --- |
| 62/200 (31.0%) | 72/200 (36.5%) | 0.2868 |

During phase 1 the experts appear to have a better OS prediction correctness. However, from the Wilcoxon signed-rank test, no statistical differences between the scores’distributions were found at the 5% level, hence no definitive conclusion can be made about this difference of the total score between experts and non-experts in phase 1.

### 4. ANALYSIS 3: EXPERTS vs NON-EXPERTS (PHASE 2)

#### 4.1 DCR

##### Table 32. Contingency table for DCR predictions by experts and non-experts in phase 2.

| **Predicted DCR by experts** | **Predicted DCR by non-experts** | |
| --- | --- | --- |
|  | **PD** | **DCR** |
| **PD** | 12 (12.0%) | 11 (11.0%) |
| **DCR** | 6 (6.0%) | 71 (71.0%) |
| **Cohen’s Kappa** | 0.4804 (moderate agreement) | |
| **McNemar’s test p-value** | 0.2253 (no significant difference between discordant predictions) | |

##### Table 33. Association between DCR prediction success and physicians’expertise in phase 2. Univariable logistic regression model.

| **N = 200** |  |  |
| --- | --- | --- |
| **Level of expertise** |  | p = 0.6576 |
| Non-expert | reference |  |
| Expert | OR (95% CI) = 0.88 (0.49-1.57) | p = 0.6576 |
| **AUC (95% CI)** | 0.516 (0.444-0.589) |  |
| **Legend:** N: number of predictions. OR: Odds Ratio. AUC: Area Under the Curve. CI: Confidence Interval. | | |

**NOTE:** ORR = 0.88 means that being an expert reduces the probability of success (i.e., predicting the true DCR) by 12% in phase 2. However, this association is not statistically significant.

##### Table 34. Metrics summary for DCR predictions according to physicians’expertise in phase 2.

|  | **Non-expert** | **Expert** | **p-value** |
| --- | --- | --- | --- |
| **Sensitivity (95% CI)** | 0.900 (0.795-0.962) | 0.833 (0.715-0.917) | 0.1573 |
| **Specificity (95% CI)** | 0.300 (0.166-0.465) | 0.325 (0.186-0.491) | 0.7389 |
| **Accuracy (95% CI)** | 0.660 (0.559-0.752) | 0.630 (0.528-0.724) | 0.4669 |
| **Precision (95% CI)** | 0.659 (0.546-0.760) | 0.649 (0.532-0.755) | 0.4142 |
| **F1-Score (95% CI)** | 0.761 (0.678-0.830) | 0.730 (0.641-0.808) | - |

After using the XAI, there are no significant differences between experts and non-experts in terms of performance.

## 4.2 OS

##### Table 35. Contingency table for OS predictions by experts and non-experts in phase 2.

| **Predicted OS by experts** | **Predicted OS by non-experts** | | | | |
| --- | --- | --- | --- | --- | --- |
|  | **< 6 months** | **[6-12) months** | **[12-18) months** | **[18-24) months** | **≥24 months** |
| **< 6 months** | 2 (2.0%) | 2 (2.0%) | 0 (0.0%) | 0 (0.0%) | 3 (3.0%) |
| **[6-12) months** | 4 (4.0%) | 12 (12.0%) | 9 (9.0%) | 1 (1.0%) | 1 (1.0%) |
| **[12-18) months** | 1 (1.0%) | 11 (11.0%) | 12 (12.0%) | 3 (3.0%) | 5 (5.0%) |
| **[18-24) months** | 0 (0.0%) | 2 (2.0%) | 9 (9.0%) | 5 (5.0%) | 5 (5.0%) |
| **≥24 months** | 0 (0.0%) | 1 (1.0%) | 3 (3.0%) | 2 (2.0%) | 7 (7.0%) |
| **Cohen’s Weighted Kappa** | 0.3359 (fair agreement) | | | | |

##### Table 36. Association between OS prediction success and physicians’expertise in phase 2. Univariable logistic regression model.

| **N = 200** |  |  |
| --- | --- | --- |
| **Level of expertise** |  | p = 0.3848 |
| Non-expert | reference |  |
| Expert | OR (95% CI) = 0.78 (0.44-1.37) | p = 0.3848 |
| **AUC (95% CI)** | 0.532 (0.460-0.603) |  |
| **Legend:** N: number of predictions. OR: Odds Ratio. AUC: Area Under the Curve. CI: Confidence Interval. | | |

**NOTE:** ORR = 0.78 means that being an expert reduces the probability of success (i.e., predicting the true OS) by 22% in phase 2. However, this association is not statistically significant.

##### Table 37. Total score for OS predictions according to physicians’expertise in phase 2.

| **Non-expert** | **Expert** | **Wilcoxon signed-rank test, p-value** |
| --- | --- | --- |
| 75/200 (37.5%) | 77/200 (38.5%) | 0.8187 |

During phase 2 the experts appear to have a slightly better OS prediction correctness. However, from the Wilcoxon signed-rank test, no statistical differences between the scores’distributions were found at the 5% level, hence no definitive conclusion can be made about this difference of the total score between experts and non-experts in phase 2.

### 5. ANALYSIS 4: CONFUSION MATRIX

##### Table 38. Confusion matrix for DCR predictions after using the XAI for all physicians.

|  | **Physician correct** |  | **Physician wrong** |  |
| --- | --- | --- | --- | --- |
| **XAI correct** | 0 (0.0%) | 85 (42.5%) | 35 (17.5%) | 12 (6.0%) |
| **XAI wrong** | 20 (10.0%) | 9 (4.5%) | 0 (0.0%) | 39 (19.5%) |
|  | *Physician changes after XAI advice* | *Physician stays after XAI advice* | *Physician changes after XAI advice* | *Physician stays after XAI advice* |

Considering the 114 cases where a physician was right, 20 physicians (17.5%) changed opinion after using the XAI and were wrong, whereas 9 (7.9%) stayed with their initial evaluation and were right.	Considering the 29 subcases where a physician was right and the XAI was wrong, 20 physicians (69.0%) changed opinion after using the XAI and were wrong, whereas 9 (31%) stayed with their initial evaluation and were right.

With regard to the 86 cases where a physician was wrong, 35 physicians (40.7%) changed their opinion after using the XAI and were right, while 12 (14.0%) stayed with their wrong evaluation.	With regard to the 47 subcases a physician was wrong and the XAI was right, 35 physicians (74.5%) changed their opinion after using the XAI and were right, while 12 (25.5%) stayed with their wrong evaluation.

##### Table 39. Confusion matrix for DCR predictions after using the XAI for expert physicians.

|  | **Physician correct** |  | **Physician wrong** |  |
| --- | --- | --- | --- | --- |
| **XAI correct** | 0 (0.0%) | 40 (40.0%) | 18 (18.0%) | 8 (8.0%) |
| **XAI wrong** | 13 (13.0%) | 5 (5.0%) | 0 (0.0%) | 16 (16.0%) |
|  | *Physician changes after XAI advice* | *Physician stays after XAI advice* | *Physician changes after XAI advice* | *Physician stays after XAI advice* |

Considering the 58 cases where an expert was right, 13 experts (22.4%) changed opinion after using the XAI and were wrong, whereas 5 (8.6%) stayed with their initial evaluation and were right. Considering the 18 subcases where an expert was right and the XAI was wrong, 13 experts (72.2%) changed opinion after using the XAI and were wrong, whereas 5 (2.8%) stayed with their initial evaluation and were right.

With regard to the 42 cases where an expert was wrong, 18 experts (42.9%) changed their opinion after using the XAI and were right, while 8 (19.0%) stayed with their wrong evaluation.	With regard to the 26 subcases a physician was wrong and the XAI was right, 18 experts (69.2%) changed their opinion after using the XAI and were right, while 8 (30.8%) stayed with their wrong evaluation.

##### Table 40. Confusion matrix for DCR predictions after using the XAI for non-expert physicians.

|  | **Physician correct** |  | **Physician wrong** |  |
| --- | --- | --- | --- | --- |
| **XAI correct** | 0 (0.0%) | 45 (45.0%) | 17 (17.0%) | 4 (4.0%) |
| **XAI wrong** | 7 (7.0%) | 4 (4.0%) | 0 (0.0%) | 23 (23.0%) |
|  | *Physician changes after XAI advice* | *Physician stays after XAI advice* | *Physician changes after XAI advice* | *Physician stays after XAI advice* |

Considering the 56 cases where a non-expert was right, 7 non-experts (12.5%) changed opinion after using the XAI and were wrong, whereas 4 (7.1%) stayed with their initial evaluation and were right. Considering the 11 subcases where a non-expert was right and the XAI was wrong, 7 non-experts (63.6%) changed opinion after using the XAI and were wrong, whereas 4 (36.4%) stayed with their initial evaluation and were right.

With regard to the 44 cases where a non-expert was wrong, 17 non-experts (38.6%) changed their opinion after using the XAI and were right, while 4 (9.1%) stayed with their wrong evaluation.	With regard to the 21 subcases a non-expert was wrong and the XAI was right, 17 non-experts (81.0%) changed their opinion after using the XAI and were right, while 4 (19.0%) stayed with their wrong evaluation.
