## Supplementary_Info_3 for "I3LUNG: Clinical Validation of a Multimodal AI Tool to Support Immunotherapy Decisions in NSCLC"

**TABLE OF CONTENTS**

**1. COHORTS 2 AND 3	2**

Table 3. Patients characteristics according to center	2

Table 4. Treatment and tumor characteristics according to center	9

### 1. COHORTS 2 AND 3

#### Table 1. Patients characteristics according to center

|  | **INT**  **N=684** | **GHD**  **N=430** | **MH**  **N=231** | **SZMC**  **N=273** | **UOC**  **N=251** | **VHIO**  **N=206** | **p-value** | **Overall N=2075** |
| --- | --- | --- | --- | --- | --- | --- | --- | --- |
| **Gender - n (%)** |  |  |  |  |  |  | <0.0001** |  |
| Male | 436 (63.7) | 262 (60.9) | 176 (76.2) | 176 (64.5) | 148 (59.0) | 138 (67.0) |  | 1291 (62.2) |
| Female | 248 (36.3) | 168 (39.1) | 55 (23.8) | 97 (35.5) | 103 (41.0) | 68 (33.0) |  | 784 (37.8) |
| **Menopausal - n (%)** |  |  |  |  |  |  | <0.0001**** |  |
| No | 17 (6.9) | 32 (19.3) | 6 (11.1) | 13 (18.1) | 6 (5.4) | 16 (23.5) |  | 90 (12.5) |
| Yes | 231 (93.1) | 134 (80.7) | 48 (88.9) | 59 (81.9) | 105 (94.6) | 52 (76.5) |  | 629 (87.5) |
| Missing | 0 | 2 | 1 | 25 | 37 | 0 |  | 65 |
| **Previous Pregnancy - n (%)** |  |  |  |  |  |  | 0.0224**** |  |
| No | 1 (6.3) | 1 (12.5) | 14 (41.2) | 1 (20.0) | 5 (17.9) | 0 (0.0) |  | 22 (21.8) |
| Yes | 15 (93.8) | 7 (87.5) | 20 (58.8) | 4 (80.0) | 23 (82.1) | 10 (100.0) |  | 79 (78.2) |
| Missing | 232 | 160 | 21 | 92 | 120 | 58 |  | 683 |
| **Age at Diagnosis (Years)** |  |  |  |  |  |  | 0.0021* |  |
| Mean (SD) | 66.3 (10.0) | 65.9 (9.5) | 66.4 (9.4) | 66.9 (9.3) | 67.0 (9.8) | 63.2 (11.5) |  | 66.1 (9.9) |
| Median (Q1 - Q3) | 67.4 (60.2-73.4) | 66.1 (59.1-73.5) | 67.8 (60.9-73.2) | 67.8 (60.5-72.8) | 67.3 (60.0-73.9) | 62.9 (55.3-71.9) |  | 66.9 (59.6-73.3) |
| Min - Max | 25.7 - 92.7 | 33.5 - 92.5 | 35.4 - 87.4 | 34.3 - 93.4 | 34.8 - 91.7 | 34.4 - 89.9 |  | 25.7 - 93.4 |
| Missing | 0 | 0 | 1 | 7 | 1 | 0 |  | 9 |
| **BMI at Baseline** |  |  |  |  |  |  | 0.0009* |  |
| Mean (SD) | 24.2 (4.1) | 24.7 (4.7) | 25.7 (4.7) | 24.5 (5.0) | 24.8 (5.4) | 25.5 (4.6) |  | 24.7 (4.7) |
| Median (Q1 - Q3) | 23.9 (21.4-26.8) | 24.4 (21.6-27.2) | 25.1 (22.9-28.1) | 23.9 (21.3-27.0) | 24.7 (20.5-28.7) | 25.5 (21.8-28.8) |  | 24.3 (21.5-27.5) |
| Min - Max | 13.5 - 46.2 | 14.4 - 45.8 | 17.5 - 59.0 | 3.7 - 37.5 | 12.6 - 42.2 | 14.7 - 38.8 |  | 3.7 - 59.0 |
| Missing | 75 | 30 | 2 | 35 | 0 | 104 |  | 246 |
| **Professional Status - n (%)** |  |  |  |  |  |  | <0.0001** |  |
| No | 3 (6.4) | 10 (21.3) | 93 (78.8) | 4 (8.5) | 69 (60.5) | 1 (6.7) |  | 180 (46.4) |
| Yes | 44 (93.6) | 37 (78.7) | 25 (21.2) | 43 (91.5) | 45 (39.5) | 14 (93.3) |  | 208 (53.6) |
| Missing | 637 | 383 | 113 | 226 | 137 | 191 |  | 1687 |
| **Level of Education - n (%)** |  |  |  |  |  |  | 0.0254**** |  |
| Degree | 6 (85.7) | 3 (100.0) | 21 (52.5) | 16 (94.1) | 28 (66.7) | 0 (0.0) |  | 74 (67.9) |
| Primary school | 0 (0.0) | 0 (0.0) | 1 (2.5) | 0 (0.0) | 0 (0.0) | 0 (0.0) |  | 1 (0.9) |
| Secondary school | 1 (14.3) | 0 (0.0) | 18 (45.0) | 1 (5.9) | 14 (33.3) | 0 (0.0) |  | 34 (31.2) |
| Missing | 677 | 427 | 191 | 256 | 209 | 206 |  | 1966 |
| **Residence - n (%)** |  |  |  |  |  |  | <0.0001**** |  |
| City | 339 (99.7) | 0 (0.0) | 177 (86.3) | 55 (98.2) | 78 (32.5) | 11 (91.7) |  | 660 (77.4) |
| Village | 1 (0.3) | 0 (0.0) | 28 (13.7) | 1 (1.8) | 162 (67.5) | 1 (8.3) |  | 193 (22.6) |
| Missing | 344 | 430 | 26 | 217 | 11 | 194 |  | 1222 |
| **Smoking Status - n (%)** |  |  |  |  |  |  | <0.0001** |  |
| Never | 50 (7.8) | 19 (5.6) | 14 (6.3) | 30 (11.8) | 23 (9.2) | 23 (11.2) |  | 159 (8.3) |
| Former | 407 (63.8) | 144 (42.4) | 95 (42.8) | 114 (44.7) | 189 (75.3) | 127 (61.7) |  | 1076 (56.3) |
| Current | 181 (28.4) | 177 (52.1) | 113 (50.9) | 111 (43.5) | 39 (15.5) | 56 (27.2) |  | 677 (35.4) |
| Missing | 46 | 90 | 9 | 18 | 2 | 0 |  | 163 |
| **Smoking Packs per Year (Among Current Smokers)** |  |  |  |  |  |  | <0.0001* |  |
| Mean (SD) | 45.8 (61.5) | 46.2 (20.0) | 67.4 (34.0) | 68.7 (36.2) | 74.1 (101.9) | 44.1 (20.4) |  | 55.1 (42.0) |
| Median (Q1 - Q3) | 37.3 (18.0-50.0) | 40.0 (35.0-59.0) | 60.0 (40.0-100.0) | 60.0 (40.0-100.0) | 40.0 (30.0-60.0) | 40.0 (30.0-50.0) |  | 50.0 (35.0-65.0) |
| Min - Max | 0.9 - 547.5 | 2.0 - 120.0 | 1.0 - 175.0 | 20.0 - 250.0 | 7.0 - 365.0 | 2.0 - 100.0 |  | 0.9 - 547.5 |
| Missing | 87 | 7 | 0 | 21 | 19 | 1 |  | 135 |
| **E-Cigarettes Use - n (%)** |  |  |  |  |  |  | 0.1631*** |  |
| No | 344 (98.3) | 14 (93.3) | 136 (97.1) | 52 (98.1) | 210 (99.5) | 1 (100.0) |  | 757 (98.3) |
| Yes | 6 (1.7) | 1 (6.7) | 4 (2.9) | 1 (1.9) | 1 (0.5) | 0 (0.0) |  | 13 (1.7) |
| Missing | 334 | 415 | 91 | 220 | 40 | 205 |  | 1305 |
| **Exposure to Passive Smoking - n (%)** |  |  |  |  |  |  | 0.0025*** |  |
| No | 318 (98.1) | 1 (100.0) | 131 (100.0) | 23 (88.5) | 161 (100.0) | 0 (0.0) |  | 634 (98.6) |
| Yes | 6 (1.9) | 0 (0.0) | 0 (0.0) | 3 (11.5) | 0 (0.0) | 0 (0.0) |  | 9 (1.4) |
| Missing | 360 | 429 | 100 | 247 | 90 | 206 |  | 1432 |
| **Level of Alcohol Consumption** |  |  |  |  |  |  | <0.0001** |  |
| Abuse | 6 (17.1) | 35 (53.8) | 27 (12.6) | 0 (0.0) | 36 (15.2) | 7 (70.0) |  | 111 (18.5) |
| No | 20 (57.1) | 10 (15.4) | 146 (68.2) | 39 (97.5) | 128 (54.0) | 1 (10.0) |  | 344 (57.2) |
| Social | 9 (25.7) | 20 (30.8) | 41 (19.2) | 1 (2.5) | 73 (30.8) | 2 (20.0) |  | 146 (24.3) |
| Missing | 649 | 365 | 17 | 233 | 14 | 196 |  | 1474 |
| **Drugs Consumption** |  |  |  |  |  |  | <0.0001**** |  |
| No | 13 (81.3) | 16 (80.0) | 136 (99.3) | 33 (89.2) | 200 (85.1) | 0 (0.0) |  | 398 (89.4) |
| Yes | 3 (18.8) | 4 (20.0) | 1 (0.7) | 4 (10.8) | 35 (14.9) | 0 (0.0) |  | 47 (10.6) |
| Missing | 668 | 410 | 94 | 236 | 16 | 206 |  | 1630 |
| **Allergies - n (%)** |  |  |  |  |  |  | <0.0001** |  |
| No | 421 (71.5) | 35 (66.0) | 217 (93.9) | 143 (65.0) | 119 (48.4) | 4 (40.0) |  | 939 (69.6) |
| Yes | 168 (28.5) | 18 (34.0) | 14 (6.1) | 77 (35.0) | 127 (51.6) | 6 (60.0) |  | 410 (30.4) |
| Missing | 95 | 377 | 0 | 53 | 5 | 196 |  | 726 |
| **Number of Allergies (Among those with allergies)** |  |  |  |  |  |  | 0.0001* |  |
| Mean (SD) | 1.5 (1.0) | 1.7 (0.9) | 1.7 (1.4) | 1.3 (0.9) | 1.9 (1.4) | 1.8 (1.3) |  | 1.6 (1.2) |
| Median (Q1 - Q3) | 1.0 (1.0-2.0) | 1.5 (1.0-2.0) | 1.0 (1.0-2.0) | 1.0 (1.0-1.0) | 1.0 (1.0-2.0) | 1.0 (1.0-3.0) |  | 1.0 (1.0-2.0) |
| Min - Max | 0.0 - 7.0 | 1.0 - 4.0 | 1.0 - 6.0 | 0.0 - 6.0 | 1.0 - 7.0 | 1.0 - 4.0 |  | 0.0 - 7.0 |
| **Oncological Comorbidities - n (%)** |  |  |  |  |  |  | <0.0001** |  |
| No | 435 (75.4) | 53 (35.3) | 198 (86.8) | 181 (85.0) | 202 (84.5) | 5 (35.7) |  | 1074 (75.6) |
| Yes | 142 (24.6) | 97 (64.7) | 30 (13.2) | 32 (15.0) | 37 (15.5) | 9 (64.3) |  | 347 (24.4) |
| Missing | 107 | 280 | 3 | 60 | 12 | 192 |  | 654 |
| **ECOG Performance Status at Baseline - n (%)** |  |  |  |  |  |  | <0.0001** |  |
| 0-1 | 567 (83.8) | 298 (79.9) | 216 (94.7) | 145 (90.6) | 203 (81.9) | 174 (87.4) |  | 1603 (85.0) |
| 2-3-4 | 110 (16.2) | 75 (20.1) | 12 (5.3) | 15 (9.4) | 45 (18.1) | 25 (12.6) |  | 282 (15.0) |
| Missing | 7 | 57 | 3 | 113 | 3 | 7 |  | 190 |
| **Concomitant Disease - n (%)** |  |  |  |  |  |  | <0.0001** |  |
| No | 76 (11.2) | 38 (9.8) | 58 (25.1) | 23 (8.5) | 32 (13.9) | 49 (23.9) |  | 276 (13.8) |
| Yes | 604 (88.8) | 351 (90.2) | 173 (74.9) | 247 (91.5) | 199 (86.1) | 156 (76.1) |  | 1730 (86.2) |
| Missing | 4 | 41 | 0 | 3 | 20 | 1 |  | 69 |
| **Cardiovascular Disorders - n (%)** |  |  |  |  |  |  | <0.0001** |  |
| No | 164 (27.2) | 117 (33.4) | 38 (22.0) | 104 (42.3) | 59 (29.6) | 56 (36.8) |  | 538 (31.2) |
| Yes | 439 (72.8) | 233 (66.6) | 135 (78.0) | 142 (57.7) | 140 (70.4) | 96 (63.2) |  | 1185 (68.8) |
| Missing | 1 | 1 | 0 | 1 | 0 | 4 |  | 7 |
| **Renal Disorders - n (%)** |  |  |  |  |  |  | <0.0001** |  |
| No | 546 (90.5) | 343 (98.0) | 168 (97.1) | 221 (89.8) | 176 (88.4) | 130 (85.5) |  | 1584 (91.9) |
| Yes | 57 (9.5) | 7 (2.0) | 5 (2.9) | 25 (10.2) | 23 (11.6) | 22 (14.5) |  | 139 (8.1) |
| Missing | 1 | 1 | 0 | 1 | 0 | 4 |  | 7 |
| **Other Neoplasms - n (%)** |  |  |  |  |  |  | <0.0001** |  |
| No | 530 (87.9) | 290 (82.9) | 165 (95.4) | 226 (91.9) | 185 (93.0) | 140 (92.1) |  | 1536 (89.1) |
| Yes | 73 (12.1) | 60 (17.1) | 8 (4.6) | 20 (8.1) | 14 (7.0) | 12 (7.9) |  | 187 (10.9) |
| Missing | 1 | 1 | 0 | 1 | 0 | 4 |  | 7 |
| **Endocrine Disorders - n (%)** |  |  |  |  |  |  | <0.0001** |  |
| No | 462 (76.6) | 341 (97.4) | 155 (89.6) | 222 (90.2) | 149 (74.9) | 144 (94.7) |  | 1473 (85.5) |
| Yes | 141 (23.4) | 9 (2.6) | 18 (10.4) | 24 (9.8) | 50 (25.1) | 8 (5.3) |  | 250 (14.5) |
| Missing | 1 | 1 | 0 | 1 | 0 | 4 |  | 7 |
| **Pulmonary Disease - n (%)** |  |  |  |  |  |  | <0.0001** |  |
| No | 467 (77.4) | 202 (57.7) | 130 (75.1) | 131 (53.3) | 141 (70.9) | 99 (65.1) |  | 1170 (67.9) |
| Yes | 136 (22.6) | 148 (42.3) | 43 (24.9) | 115 (46.7) | 58 (29.1) | 53 (34.9) |  | 553 (32.1) |
| Missing | 1 | 1 | 0 | 1 | 0 | 4 |  | 7 |
| **Autoimmune Disorders - n (%)** |  |  |  |  |  |  | <0.0001** |  |
| No | 537 (89.1) | 335 (95.7) | 169 (97.7) | 213 (86.6) | 158 (79.4) | 146 (96.1) |  | 1558 (90.4) |
| Yes | 66 (10.9) | 15 (4.3) | 4 (2.3) | 33 (13.4) | 41 (20.6) | 6 (3.9) |  | 165 (9.6) |
| Missing | 1 | 1 | 0 | 1 | 0 | 4 |  | 7 |
| **Hepatic Disorders - n (%)** |  |  |  |  |  |  | <0.0001** |  |
| No | 554 (91.9) | 342 (97.7) | 172 (99.4) | 228 (92.7) | 180 (90.5) | 145 (95.4) |  | 1621 (94.1) |
| Yes | 49 (8.1) | 8 (2.3) | 1 (0.6) | 18 (7.3) | 19 (9.5) | 7 (4.6) |  | 102 (5.9) |
| Missing | 1 | 1 | 0 | 1 | 0 | 4 |  | 7 |
| **Infections - n (%)** |  |  |  |  |  |  | <0.0001** |  |
| No | 490 (81.3) | 341 (97.4) | 170 (98.3) | 200 (81.3) | 137 (68.8) | 145 (95.4) |  | 1483 (86.1) |
| Yes | 113 (18.7) | 9 (2.6) | 3 (1.7) | 46 (18.7) | 62 (31.2) | 7 (4.6) |  | 240 (13.9) |
| Missing | 1 | 1 | 0 | 1 | 0 | 4 |  | 7 |
| **Gastrointestinal Disorders - n (%)** |  |  |  |  |  |  | <0.0001** |  |
| No | 513 (85.1) | 342 (97.7) | 167 (96.5) | 218 (88.6) | 168 (84.4) | 129 (84.9) |  | 1537 (89.2) |
| Yes | 90 (14.9) | 8 (2.3) | 6 (3.5) | 28 (11.4) | 31 (15.6) | 23 (15.1) |  | 186 (10.8) |
| Missing | 1 | 1 | 0 | 1 | 0 | 4 |  | 7 |
| **Diabetes - n (%)** |  |  |  |  |  |  | <0.0001** |  |
| No | 527 (87.4) | 287 (82.0) | 127 (73.4) | 182 (74.0) | 156 (78.4) | 119 (78.3) |  | 1398 (81.1) |
| Yes | 76 (12.6) | 63 (18.0) | 46 (26.6) | 64 (26.0) | 43 (21.6) | 33 (21.7) |  | 325 (18.9) |
| Missing | 1 | 1 | 0 | 1 | 0 | 4 |  | 7 |
| **Neurological Disorders - n (%)** |  |  |  |  |  |  | <0.0001** |  |
| No | 548 (90.9) | 343 (98.0) | 168 (97.1) | 228 (92.7) | 179 (89.9) | 148 (97.4) |  | 1614 (93.7) |
| Yes | 55 (9.1) | 7 (2.0) | 5 (2.9) | 18 (7.3) | 20 (10.1) | 4 (2.6) |  | 109 (6.3) |
| Missing | 1 | 1 | 0 | 1 | 0 | 4 |  | 7 |
| **Dislipidemia - n (%)** |  |  |  |  |  |  | <0.0001** |  |
| No | 497 (82.4) | 341 (97.4) | 108 (62.4) | 201 (81.7) | 124 (62.3) | 84 (55.3) |  | 1355 (78.6) |
| Yes | 106 (17.6) | 9 (2.6) | 65 (37.6) | 45 (18.3) | 75 (37.7) | 68 (44.7) |  | 368 (21.4) |
| Missing | 1 | 1 | 0 | 1 | 0 | 4 |  | 7 |
| **Legend:** N: number of subjects. SD: Standard Deviation. Q1 - Q3: First - Third Quartile. Min - Max: Minimum - Maximum  *: p-value of Kruskal-Wallis’ Test for Medians **: p-value of Chi-Square Test ***: p-value of Fisher’s Exact Test ****: p-value of Fisher’s Exact Test based on 10000 Monte Carlo simulations | | | | | | | | |

#### Table 2. Treatment and tumor characteristics according to center

|  | **INT**  **N=684** | **GHD**  **N=430** | **MH**  **N=231** | **SZMC**  **N=273** | **UOC**  **N=251** | **VHIO**  **N=206** | **p-value** | **Overall N=2075** |
| --- | --- | --- | --- | --- | --- | --- | --- | --- |
| **Treatment Arm (Immuno Alone vs Immuno + Chemo vs Immuno + Other) - n (%)** |  |  |  |  |  |  | <0.0001**** |  |
| Immuno Alone | 250 (55.9) | 126 (46.3) | 33 (17.0) | 40 (17.4) | 72 (34.1) | 52 (61.9) |  | 573 (39.8) |
| Immuno + Chemo | 197 (44.1) | 140 (51.5) | 160 (82.5) | 189 (82.2) | 139 (65.9) | 32 (38.1) |  | 857 (59.6) |
| Immuno + Other | 0 (0.0) | 6 (2.2) | 1 (0.5) | 1 (0.4) | 0 (0.0) | 0 (0.0) |  | 8 (0.6) |
| Missing | 237 | 158 | 37 | 43 | 40 | 122 |  | 637 |
| **Treatment Arm (Immuno Alone/Immuno + Other**  **vs Immuno + Chemo) - n (%)** |  |  |  |  |  |  | <0.0001** |  |
| Immuno Alone/Immuno + Other | 250 (55.9) | 132 (48.5) | 34 (17.5) | 41 (17.8) | 72 (34.1) | 52 (61.9) |  | 581 (40.4) |
| Immuno + Chemo | 197 (44.1) | 140 (51.5) | 160 (82.5) | 189 (82.2) | 139 (65.9) | 32 (38.1) |  | 857 (59.6) |
| Missing | 237 | 158 | 37 | 43 | 40 | 122 |  | 637 |
| **Immunotherapy line - n (%)** |  |  |  |  |  |  | <0.0001** |  |
| 1st | 447 (65.4) | 272 (63.3) | 194 (84.0) | 230 (84.2) | 210 (83.7) | 84 (40.8) |  | 1437 (69.3) |
| 2nd | 149 (21.8) | 107 (24.9) | 31 (13.4) | 39 (14.3) | 36 (14.3) | 84 (40.8) |  | 446 (21.5) |
| 3rd or further | 88 (12.9) | 51 (11.9) | 6 (2.6) | 4 (1.5) | 5 (2.0) | 38 (18.4) |  | 192 (9.3) |
| **Age at Treatment Start (Years)** |  |  |  |  |  |  | 0.0025* |  |
| Mean (SD) | 67.4 (9.9) | 66.7 (9.6) | 67.5 (9.4) | 67.2 (9.1) | 68.4 (10.0) | 64.4 (11.3) |  | 67.0 (9.9) |
| Median (Q1 - Q3) | 68.6 (61.2-74.3) | 67.1 (60.1-74.3) | 68.8 (61.4-73.9) | 68.1 (61.9-73.2) | 69.3 (61.8-75.6) | 64.7 (57.2-72.6) |  | 67.9 (60.5-74.1) |
| Min - Max | 27.5 - 92.8 | 34.1 - 92.6 | 35.6 - 87.0 | 34.4 - 93.5 | 35.9 - 91.7 | 34.6 - 90.0 |  | 27.5 - 93.5 |
| Missing | 4 | 13 | 22 | 63 | 125 | 0 |  | 227 |
| **Histology (Non Squamous vs Squamous) - n (%)** |  |  |  |  |  |  | <0.0001** |  |
| Non Squamous | 551 (81.0) | 293 (68.3) | 166 (72.5) | 195 (73.9) | 220 (89.1) | 163 (80.3) |  | 1588 (77.4) |
| Squamous | 129 (19.0) | 136 (31.7) | 63 (27.5) | 69 (26.1) | 27 (10.9) | 40 (19.7) |  | 464 (22.6) |
| Missing | 4 | 1 | 2 | 9 | 4 | 3 |  | 23 |
| **Biopsy Type - n (%)** |  |  |  |  |  |  | <0.0001** |  |
| Histological | 641 (94.8) | 140 (42.0) | 185 (80.1) | 207 (95.4) | 43 (17.2) | 10 (100.0) |  | 1226 (71.4) |
| Citological | 22 (3.3) | 103 (30.9) | 14 (6.1) | 5 (2.3) | 166 (66.4) | 0 (0.0) |  | 310 (18.1) |
| Surgery Specimen | 13 (1.9) | 90 (27.0) | 32 (13.9) | 5 (2.3) | 41 (16.4) | 0 (0.0) |  | 181 (10.5) |
| Missing | 8 | 97 | 0 | 56 | 1 | 196 |  | 358 |
| **Biopsy Site (Lung vs Other) - n (%)** |  |  |  |  |  |  | <0.0001** |  |
| Lung | 420 (63.2) | 275 (84.6) | 186 (80.5) | 194 (86.6) | 135 (54.0) | 5 (50.0) |  | 1215 (71.3) |
| Other | 245 (36.8) | 50 (15.4) | 45 (19.5) | 30 (13.4) | 115 (46.0) | 5 (50.0) |  | 490 (28.7) |
| Missing | 19 | 105 | 0 | 49 | 1 | 196 |  | 370 |
| **PDL1 Percentage - n (%)** |  |  |  |  |  |  | <0.0001** |  |
| < 1% | 198 (33.3) | 69 (24.0) | 90 (42.5) | 83 (36.6) | 101 (45.9) | 63 (48.8) |  | 604 (36.2) |
| 1-49% | 219 (36.9) | 101 (35.1) | 73 (34.4) | 41 (18.1) | 50 (22.7) | 23 (17.8) |  | 507 (30.4) |
| ≥ 50% | 177 (29.8) | 118 (41.0) | 49 (23.1) | 103 (45.4) | 69 (31.4) | 43 (33.3) |  | 559 (33.5) |
| Missing | 90 | 142 | 19 | 46 | 31 | 77 |  | 405 |
| **Number of Metastases (1 vs > 1) - n (%)** |  |  |  |  |  |  | 0.0009** |  |
| 1 | 315 (49.8) | 155 (50.5) | 132 (67.0) | 130 (56.0) | 105 (49.5) | 107 (52.5) |  | 944 (52.9) |
| > 1 | 317 (50.2) | 152 (49.5) | 65 (33.0) | 102 (44.0) | 107 (50.5) | 97 (47.5) |  | 840 (47.1) |
| Missing | 52 | 123 | 34 | 41 | 39 | 2 |  | 291 |
| **Number of Metastases** |  |  |  |  |  |  | <0.0001* |  |
| Mean (SD) | 1.8 (1.1) | 1.8 (1.1) | 1.4 (0.7) | 1.7 (1.0) | 1.8 (1.0) | 1.9 (1.3) |  | 1.8 (1.1) |
| Median (Q1 - Q3) | 2.0 (1.0-2.0) | 1.0 (1.0-2.0) | 1.0 (1.0-2.0) | 1.0 (1.0-2.0) | 2.0 (1.0-2.0) | 1.0 (1.0-2.0) |  | 1.0 (1.0-2.0) |
| Min - Max | 1.0 - 7.0 | 1.0 - 7.0 | 1.0 - 4.0 | 1.0 - 7.0 | 1.0 - 6.0 | 1.0 - 7.0 |  | 1.0 - 7.0 |
| Missing | 52 | 123 | 34 | 41 | 39 | 2 |  | 291 |
| **Diagnosis Stage (All stages) - n (%)** |  |  |  |  |  |  | <0.0001**** |  |
| I | 36 (5.3) | 2 (0.5) | 12 (5.2) | 2 (0.7) | 11 (4.4) | 11 (5.4) |  | 74 (3.6) |
| II | 35 (5.2) | 3 (0.7) | 7 (3.1) | 8 (3.0) | 16 (6.4) | 5 (2.5) |  | 74 (3.6) |
| III | 127 (18.9) | 128 (29.9) | 54 (23.6) | 23 (8.6) | 62 (24.7) | 41 (20.3) |  | 435 (21.2) |
| IV | 475 (70.6) | 295 (68.9) | 156 (68.1) | 235 (87.7) | 162 (64.5) | 145 (71.8) |  | 1468 (71.6) |
| Missing | 11 | 2 | 2 | 5 | 0 | 4 |  | 24 |
| **T Stage - n (%)** |  |  |  |  |  |  | <0.0001** |  |
| T0+T1+T2 | 241 (35.9) | 119 (27.7) | 73 (31.9) | 162 (61.6) | 213 (85.2) | 70 (45.5) |  | 878 (44.0) |
| T3+T4 | 430 (64.1) | 311 (72.3) | 156 (68.1) | 101 (38.4) | 37 (14.8) | 84 (54.5) |  | 1119 (56.0) |
| Missing | 13 | 0 | 2 | 10 | 1 | 52 |  | 78 |
| **N Stage - n (%)** |  |  |  |  |  |  | 0.0006** |  |
| N0+N1 | 176 (26.2) | 113 (26.3) | 75 (32.8) | 101 (38.4) | 89 (35.6) | 53 (34.2) |  | 607 (30.4) |
| N2+N3 | 495 (73.8) | 317 (73.7) | 154 (67.2) | 162 (61.6) | 161 (64.4) | 102 (65.8) |  | 1391 (69.6) |
| Missing | 13 | 0 | 2 | 10 | 1 | 51 |  | 77 |
| **M Stage - n (%)** |  |  |  |  |  |  | <0.0001** |  |
| M0 | 199 (29.6) | 131 (30.8) | 73 (31.9) | 30 (11.5) | 90 (36.1) | 60 (29.3) |  | 583 (28.6) |
| M1 | 474 (70.4) | 294 (69.2) | 156 (68.1) | 230 (88.5) | 159 (63.9) | 145 (70.7) |  | 1458 (71.4) |
| Missing | 11 | 5 | 2 | 13 | 2 | 1 |  | 34 |
| **Surgery - n (%)** |  |  |  |  |  |  | 0.7903** |  |
| No | 553 (80.8) | 345 (80.2) | 186 (81.2) | 216 (82.4) | 205 (81.7) | 159 (77.2) |  | 1664 (80.7) |
| Yes | 131 (19.2) | 85 (19.8) | 43 (18.8) | 46 (17.6) | 46 (18.3) | 47 (22.8) |  | 398 (19.3) |
| Missing | 0 | 0 | 2 | 11 | 0 | 0 |  | 13 |
| **Neoadjuvant Treatment - n (%)** |  |  |  |  |  |  | <0.0001** |  |
| No | 648 (94.9) | 44 (68.8) | 213 (93.0) | 214 (97.3) | 6 (2.7) | 3 (21.4) |  | 1128 (78.8) |
| Yes | 35 (5.1) | 20 (31.3) | 16 (7.0) | 6 (2.7) | 216 (97.3) | 11 (78.6) |  | 304 (21.2) |
| Missing | 1 | 366 | 2 | 53 | 29 | 192 |  | 643 |
| **Immunotherapy - n (%)** |  |  |  |  |  |  | <0.0001**** |  |
| No | 34 (97.1) | 19 (95.0) | 16 (100.0) | 6 (100.0) | 4 (1.9) | 11 (100.0) |  | 90 (29.6) |
| Yes | 1 (2.9) | 1 (5.0) | 0 (0.0) | 0 (0.0) | 212 (98.1) | 0 (0.0) |  | 214 (70.4) |
| **Platinum Based Chemotherapy - n (%)** |  |  |  |  |  |  | 0.0057**** |  |
| No | 2 (5.7) | 1 (5.0) | 1 (6.3) | 2 (33.3) | 56 (25.9) | 1 (9.1) |  | 63 (20.7) |
| Yes | 33 (94.3) | 19 (95.0) | 15 (93.8) | 4 (66.7) | 160 (74.1) | 10 (90.9) |  | 241 (79.3) |
| **Target Therapy - n (%)** |  |  |  |  |  |  | 0.1982**** |  |
| No | 35 (100.0) | 20 (100.0) | 15 (93.8) | 5 (83.3) | 211 (97.7) | 11 (100.0) |  | 297 (97.7) |
| Yes | 0 (0.0) | 0 (0.0) | 1 (6.3) | 1 (16.7) | 5 (2.3) | 0 (0.0) |  | 7 (2.3) |
| **Adjuvant Therapy - n (%)** |  |  |  |  |  |  | 0.0002** |  |
| No | 642 (93.9) | 395 (91.9) | 207 (89.6) | 249 (94.7) | 214 (85.3) | 181 (88.3) |  | 1888 (91.5) |
| Yes | 42 (6.1) | 35 (8.1) | 24 (10.4) | 14 (5.3) | 37 (14.7) | 24 (11.7) |  | 176 (8.5) |
| Missing | 0 | 0 | 0 | 10 | 0 | 1 |  | 11 |
| **Immunotherapy - n (%)** |  |  |  |  |  |  | <0.0001**** |  |
| No | 41 (97.6) | 33 (94.3) | 22 (91.7) | 8 (57.1) | 15 (40.5) | 24 (100.0) |  | 143 (81.3) |
| Yes | 1 (2.4) | 2 (5.7) | 2 (8.3) | 6 (42.9) | 22 (59.5) | 0 (0.0) |  | 33 (18.8) |
| **Platinum Based Chemotherapy - n (%)** |  |  |  |  |  |  | <0.0001**** |  |
| No | 1 (2.4) | 7 (20.0) | 1 (4.2) | 1 (7.1) | 14 (37.8) | 13 (54.2) |  | 37 (21.0) |
| Yes | 41 (97.6) | 28 (80.0) | 23 (95.8) | 13 (92.9) | 23 (62.2) | 11 (45.8) |  | 139 (79.0) |
| **Target Therapy - n (%)** |  |  |  |  |  |  | 0.0606**** |  |
| No | 42 (100.0) | 35 (100.0) | 24 (100.0) | 14 (100.0) | 34 (91.9) | 24 (100.0) |  | 173 (98.3) |
| Yes | 0 (0.0) | 0 (0.0) | 0 (0.0) | 0 (0.0) | 3 (8.1) | 0 (0.0) |  | 3 (1.7) |
| **Radiotherapy - n (%)** |  |  |  |  |  |  | <0.0001**** |  |
| No | 28 (66.7) | 2 (8.7) | 16 (66.7) | 6 (66.7) | 23 (63.9) | 2 (16.7) |  | 77 (52.7) |
| Yes | 14 (33.3) | 21 (91.3) | 8 (33.3) | 3 (33.3) | 13 (36.1) | 10 (83.3) |  | 69 (47.3) |
| Missing | 0 | 12 | 0 | 5 | 1 | 12 |  | 30 |
| **Legend:** N: number of subjects. SD: Standard Deviation. Q1 - Q3: First - Third Quartile. Min - Max: Minimum - Maximum.  *: p-value of Kruskal-Wallis’ Test for Medians **: p-value of Chi-Square Test ***: p-value of Fisher’s Exact Test ****: p-value of Fisher’s Exact Test based on 10000 Monte Carlo simulations | | | | | | | | |
